# Aldolase B and Angiotensinogen are associated with weekly chronic multisite spinal pain in men

**DOI:** 10.1101/2024.11.07.24316891

**Authors:** Hilla Ruotsalainen, Gabin Drouard, Andreas Pedersen, Minna Ståhl, Katja M. Kanninen, Jaakko Kaprio

## Abstract

Spinal pain problems increase with age, but already 21% of Finnish young adults suffer from a musculoskeletal disorder. Chronic widespread pain (CWP) significantly increases one’s disability level and up to 50% of people experiencing chronic pain report symptoms of depression. Diagnosing pain is challenging, as it is a subjective feeling and current clinical pain descriptors are not accurate enough to determine pain perception. In this study, variation in the human plasma proteome was investigated with untargeted Mass Spectrometry in young adult twins with weekly chronic multisite spinal pain (n=94), twins with weekly chronic local neck pain (n=99), and healthy twin individuals (n=236). The association of depression with pain and sex-specific proteins in the studied associations was also investigated. The main data analysis approach included multiple regressions which were done with Generalized Linear Mixed Models (GLMM). Of the 411 studied proteins, Aldolase B and Angiotensinogen were negatively associated with weekly chronic concomitant neck and back pain in men. These proteins are known to relate to muscle atrophy and might thus contribute to the development of musculoskeletal pain. In addition, individuals with weekly chronic concomitant neck and back pain had more symptoms of depression than individuals with weekly chronic local neck pain. Further research is needed to identify the key proteins for clinical settings and upcoming pain proteomics research should include both women and men to examine sex differences.

## Introduction

Chronic pain in young adults remains an underexplored area of research despite its considerable prevalence with more than every tenth young adult reporting chronic pain worldwide. Chronic pain impacts the quality of life especially during young adulthood, possibly affecting educational and career decisions (Murray *et al*, 2022). While older adults represent the largest group affected by pain, chronic pain often originates in childhood and the prevalence increases with age (King *et al*, 2011), affecting both physical and social well-being (Tutelman *et al*, 2021).

In a Global Burden of Disease (GBD) analysis, approximately 1.71 billion people suffered from musculoskeletal conditions in the world in 2019 (Cieza *et al*, 2020), while approximately 21% of young adults (age 20–24) in Finland experience these conditions (WHO Rehabilitation need estimator, 2019). Neck pain is the most reported persistent musculoskeletal pain among Finnish school-age children (Mikkelsson *et al*, 1997; El-Metwally *et al*, 2004). Ståhl et al (2014) studied the secular trends of single and concomitant neck and low back pain from 1990 to 2017 among 12-18-year-old Finns. They found a 2–3-fold increase in prevalence of concomitant neck and low back pain while single neck and back pain remained stable over the three decades.

People with musculoskeletal disorders are at a higher risk of developing mental health issues (WHO Musculoskeletal health, 2022). In fact, up to 50% of patients with chronic pain report major depressive disorder (Humo *et al*, 2019). Additionally, pain conditions are commonly associated with obesity in adults (Seaman, 2013; Johnston *et al*, 2019).

Chronic pain is defined as pain lasting at least three months (Raja *et al*, 2020). Rather than occurring in one anatomical location, it often manifests as multisite or widespread (Landmark *et al*, 2019). According to the ICD-11 classification, chronic widespread pain (CWP) “is diffuse musculoskeletal pain in at least 4 of 5 body regions” and “is associated with significant emotional distress (anxiety, anger/frustration, or depressed mood) or functional disability” (Nicholas *et al*, 2019). Diagnostic tools for chronic pain, such as pain drawing, visual analog, numerical rating, and verbal rating scales, rely heavily on subjective patient reporting, limiting their reliability and sensitivity (Tracey *et al*, 2019). Hence, there is a pressing need for more objective methods to complement the assessment of pain.

Blood contains valuable molecular information that could be utilized to develop biomarkers for chronic pain diagnosis and management (Yurkovich & Hood, 2019). Proteomics, the study of proteins in biological systems, offers a promising avenue for identifying biomarkers associated with chronic pain conditions (Cui *et al*, 2022).

However, research in pain proteomics, particularly focusing on CWP, is limited. Gerdle and his research group have made notable contributions (Gerdle *et al*, 2017; Stensson *et al*, 2017; Wåhlén *et al*, 2017, 2018; Gerdle *et al*, 2020, 2022). Furthermore, a recent comprehensive UK Biobank study reveals distinct proteomic signatures for various CWP types (Chen *et al*, 2024). Nevertheless, proteomic studies often lack consensus on methodologies, which makes it difficult to gather the key proteins for clinical settings in a standard fashion. Additionally, current human CWP studies predominantly involve female participants, despite evidence of sex differences in CWP prevalence (Andrews *et al*, 2018). For instance, a UK Biobank study in 2021 revealed genetic sex differences in patients with multisite chronic pain (Johnston *et al*, 2021).

This study aims to discover new protein markers for chronic spinal pain at multiple sites in the cervical, thoracic and lumbar spine, i.e. a multisite pain. We used untargeted MS-based plasma proteomics to investigate which proteins were differentially represented in Finnish young adults (FinnTwin12 cohort) with weekly chronic concomitant neck and back pain (multisite pain) compared to young adults with weekly chronic local neck pain and pain-free controls. Furthermore, this study examined the association between depressive symptoms and chronic pain and investigated sex-specific proteins in women and men with chronic multisite spinal pain.

## Methods

### Participants

Twins born 1983–1987 were 11–12 years old as they were enrolled in the FinnTwin12 (FT12) cohort through Finland’s Central Population Registry (wave 1) (Rose *et al*, 2019). In the present case-control analysis, we used 786 twins who participated in an in-person assessment as part of wave 4. These young adults (age-range 19 to 24 years) provided blood plasma samples after an overnight fast (for details see Drouard et al, 2023). Finally, 733 of those also answered health, pain, and depressive symptom (GBI) questionnaires.

All the twins provided written informed consent before the start of the study. Ethical approval for data collection was obtained from the ethical committee of the Helsinki and Uusimaa University Hospital District and Indiana University’s Institutional Review Board.

### Measurements

The wave 4 data was collected between 2006-2009 when twins were young adults. The assessment period included self-reported questionnaires and in-person interviews (psychiatric factors, anthropometrics, and neuropsychological tests) in Helsinki (Rose *et al*, 2019). The mean and variance of the group variables can be seen in Table 1. Venous blood samples were collected, and plasma was extracted within 30 minutes and stored at −80 °C until proteomic analysis in 2022.

**Table 1.** Characteristics of participants by pain group.

| Group variables | Weekly spinal CMP, n= 94 |  | Weekly chronic neck pain, n= 99 |  | Controls (pain-free), n= 236 |  |
| --- | --- | --- | --- | --- | --- | --- |
|  | Mean (min-max) | Sd | Mean (min-max) | Sd | Mean (min-max) | Sd |
| **Age (years)** | 22.2 (21.1–24.7) | 0.60 | 22.4 (21.1–24.5) | 0.64 | 22.3 (19.6–23.8) | 0.57 |
| **GBI** | 16.0 (10–35) | 5.0 | 14.3 (10–27) | 4.0 | 13.4 (10–33) | 4.0 |
| **Height (cm)** | 171.1 (149–200) | 10.5 | 170.4 (153–190) | 9.1 | 172.6 (150–196) | 9.2 |
| **Weight (kg)** | 66.2 (44–102) | 13.3 | 67.2 (48–105) | 12.5 | 70.2 (46–150) | 15.5 |
| **BMI (kg/m<sup>2</sup>)** | 22.5 (17.2–33.6) | 3.1 | 23.1 (17.9–35.1) | 3.3 | 23.4 (17.3–42) | 3.8 |
| **Males/Females** | 31/63 | NA | 32/67 | NA | 116/120 | NA |

BMI = Body Mass Index, CMP = Chronic multisite pain, GBI = General Behavior Inventory, Sd = Standard deviation

#### Pain characteristics

In this study, we use the term “chronic multisite pain” (CMP) instead of “CWP” because our pain measurements did not fully align with the criteria for CWP.

The frequency of the pain in the last three months was assessed with a 5-point Likert scale with answer options of: 1 = “almost daily”, 2 = “more than once a week”, 3 = “once a week”, 4 = “once a month”, and 5 = “rarely or never” (Appendix 1). The following anatomical regions were registered: neck, shoulders, upper and lower limbs, chest, upper (thoracic) and lower (lumbar) back, and buttocks. In the study, Likert points 1–3 were considered at least weekly pain and 4–5 no weekly pain.

Based on the pain-related questions and research done by Ståhl et al. (2014), the following four groups of pain variables were created: weekly chronic multisite spinal pain (wCMP), weekly chronic local neck pain (NECK), healthy controls with no weekly pain conditions (controls), and others (excluded from the analysis). The inclusion criterion in the wCMP group was concomitant weekly chronic pain in neck, upper, and lower back area and no other weekly pain conditions. The NECK group included those with weekly chronic neck pain and no weekly pain in other regions. The inclusion criteria in the controls group were no weekly pain conditions and considered pain-free. There were no specific exclusion criteria since all the twins were considered healthy with no major disabling conditions. All in all, 94 twins were assigned to wCMP, 99 to NECK, and 236 to controls group.

#### Psychological factors

Depressive symptoms were studied using a 10-item subscale of the GBI (Depue *et al*, 1989, Heinonen-Guzejev *et al*, 2023), with each item answered on a 4-point Likert scale with answer options of: 1 = “never”, 2 = “sometimes”, 3 = “often”, and 4 = “very often” (Appendix 2). The total sum score ranges from 10 to 40. Psychometric properties and validation against major depression diagnosis of the short GBI are given by Heinonen-Gusejev et al (2023).

#### Body Mass Index (BMI)

BMI values (kg/m^2^) were calculated from self-reported weight and height measures in the wave 4. Self-reported values were used since they were measured at the same time point as other measures and the used pain variable was also self-reported in a questionnaire that the participants completed immediately before the clinic visit. All participants in the target groups did not report BMI measures. More precisely, four, three, and four individuals did not report weight and/or height in the wCMP, NECK, and controls groups, respectively. The reliability of self-reported measures was confirmed with in-person weight and height measurements taken on the clinic visit day. As reported elsewhere, self-reported weight and height correlated strongly with measured values (Pearson correlation for weight: r=0.98 and height: r=0.99) (Drouard *et al*, 2023).

#### Preprocessing of phenotype data

The first phenotype dataset included a pain variable consisting of individuals with weekly spinal CMP and controls, and another phenotype dataset consisted of individuals with weekly chronic local neck pain and controls. Other variables in phenotype datasets were sex, age, GBI, and BMI. Preprocessing of the phenotype data at the University of Helsinki was conducted with the SPSS (version 27) and the R software (version 4.2.2).

### Proteomic data processing

#### Protein identification and quantification of protein abundances

The proteins of 786 human depleted plasma samples were first precipitated, digested, and later analyzed by a DIA method with Liquid Chromatography Electrospray Ionization Tandem Mass Spectrometry (LC-ESI-MS/MS) in 2022 at the Turku Proteomics Facility supported by Biocenter Finland. More details about high abundant protein depletion, precipitation, and digestion are available (Afonin *et al*, 2023). For DIA analysis 500 ng peptides were analyzed in a random order and analysis was performed on a nanoflow HPLC system (Easy-nLC1000, Thermo Fisher Scientific) coupled to the Q Exactive HF mass spectrometer (Thermo Fisher Scientific, Bremen, Germany) equipped with a nano-electrospray ionization source. Mass spectrometry (MS) data was obtained by using Thermo Xcalibur 4.1 software (Thermo Fisher Scientific). Data was further analyzed by Spectronaut software (Biognosys; version 16.0.2) and MaxLFQ was used to determine the label-free quantifications. Data was local-normalized. Sample IDs were matched with the questionnaire’s IDs later at FIMM, University of Helsinki.

#### Preprocessing of proteomic data

Quality of the data was controlled as described elsewhere (Drouard *et al*, 2023). Raw matrix counts were processed at the University of Helsinki. The initial proteomic data had four batches, and the batch effects were corrected using ComBat (Johnson *et al*, 2007). Protein quantities obtained from the MS analysis were log2-transformed to reduce skewness in original distributions. Age variable was scaled so that one unit correlated with one standard deviation (sd) and zero mean. As most of the values in the dataset were expected to be Missing Not at Random (NMAR), we used a special non-standard imputation method to impute missing values by the Sample minimum method (Liu & Dongre, 2021). There were no individuals for whom proteomic collection had failed, and missing value imputation was done at the proteomic level in proteins having less than 5% of missing values. Variables with more than 5% of missing values were excluded. Thus, 411 proteins were kept for further analyses. All proteomic preprocessing at the University of Helsinki was done using the R software version 4.2.2.

### Statistical analyses

We assessed associations between pain and protein levels using generalized linear mixed-effects models (GLMM) and in particular, mixed effects logistic regression as our outcomes were binary. Pain was used as an outcome and proteins were used as predictor variables in all the analyses. Fixed effect covariates included age and sex (initial model) and later also GBI and BMI in addition (fully adjusted model). Twins’ familial relatedness was adjusted for models by adding family identifiers as a random intercept. The parameters in the GLMM were estimated by restricted maximum likelihood estimation. Since we fitted hundreds of models, convergence issues were to be expected in some cases. We conducted a sensitivity analysis using Generalized Estimating Equation (GEE) models to ensure that the estimates for which convergence problems were found with GLMM were robust.

At first, GLMM and sensitivity GEE models were run to identify proteins to be associated with wCMP or NECK. Then the wCMP group was divided into two groups based on sex and initial and fully adjusted regressions were run again for both females and males to examine sex-specific effects on multisite chronic pain. Obtained nominal p-values were corrected for multiple testing with Holm method (Holm, 1979). Regressions and visualization of the results were conducted with R-studio by the way of the lme4 (version 1.1-31) and geepack (version 1.3.9) packages (R version 4.2.2).

## Results

The mean age was 22 years in all groups. GBI sum score was highest in the group with wCMP and lowest in the controls (Table 1). BMI measures ranged between 22.5 and 23.4 (kg/m^2^) with the wCMP group having the lowest and the controls having the highest mean values. Females were overrepresented in every group compared to males. Both wCMP and NECK groups included approximately 2/3 of females and 1/3 of males while sex distribution in the controls was more equal (Table 1).

### Depression symptoms were associated with the wCMP but not with the NECK group

The group with weekly spinal CMP had more depressive symptoms than controls (p-value 0.00083, age and sex-adjusted odds ratio was 1.12 [95 % Confidence Interval: 1.05,1.21]). No association was seen for the NECK group compared to controls (p-value 0.155).

### Proteins were not associated with weekly spinal CMP or weekly chronic local neck pain in initial or fully adjusted regression models

Multiple regressions were conducted using mixed-effects models. The initial and fully adjusted regression models were used to investigate plasma protein association in weekly spinal CMP and weekly chronic local neck pain. None of the plasma proteins in the wCMP or the NECK in initial or fully adjusted regression models that included all the twins remained significant after Holm correction of the p-values (summary statistics of the fully adjusted model can be found in Appendix 3 & 4).

### Angiotensinogen and Fructose-bisphosphate aldolase B were negatively associated with weekly chronic multisite spinal pain in males

An analysis stratifying by sex was performed in the wCMP group, given prior knowledge of differences in pain and covariate prevalences by sex. None of the plasma proteins were significant in females in initial or fully adjusted regressions after correcting for multiple testing. Among males, two proteins were found to be negatively associated with wCMP in fully adjusted regression (Table 2). The proteins were Angiotensinogen (AGT, protein ID: P01019) and Fructose-bisphosphate aldolase B (Aldolase B, protein ID: P05062). In boxplots, AGT and Aldolase B levels were significantly lower in men with weekly spinal CMP compared to male controls (Fig. 1A & 1B). No other plasma proteins were found when using adjustment with age and sex only.

**Table 2.**
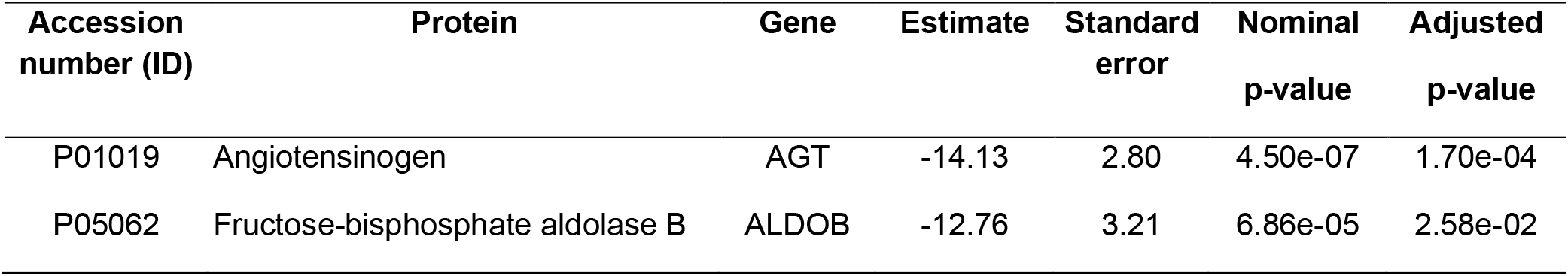
The plasma proteins associated with weekly spinal CMP in males. The fully adjusted GLM regression model included age, sex, GBI, and BMI. Aldolase B and Angiotensinogen remained significant after Holm correction.

**Figure 1.**
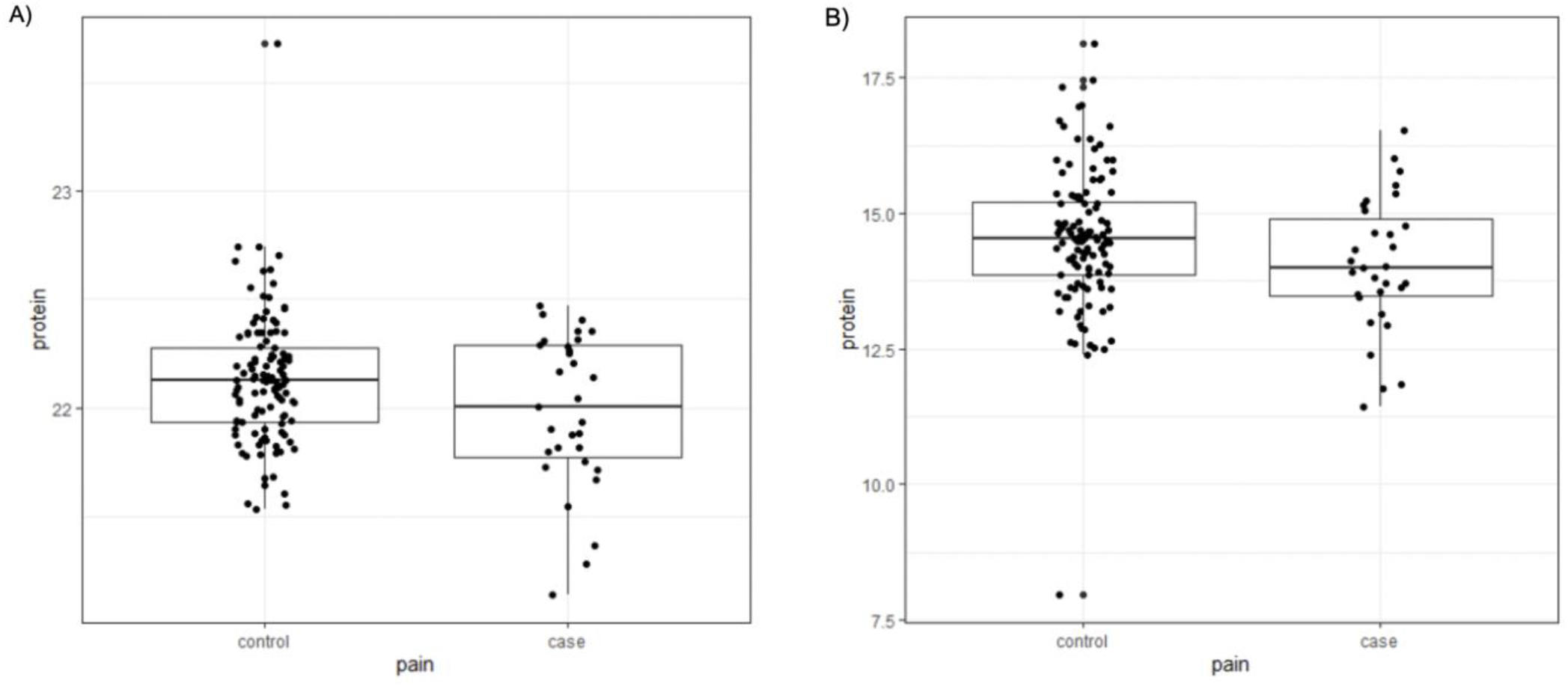
Angiotensinogen (A) and Fructose-bisphosphate aldolase B (B) levels in males with weekly spinal CMP (case) and males without pain (pain-free control). Protein levels are log2-transformed and normalized abundance values obtained from the MS analysis. Protein levels are significantly lower in the cases compared to controls.

## Discussion

In this study, the association of 411 proteins with two pain conditions in young adults was investigated. None of the proteins in initial or fully adjusted regressions (wCMP and NECK) that included all the twin individuals remained significant after correcting for multiple comparisons. Further analyses were conducted to study the effect of depression in plasma proteomes and to find sex-specific proteins in spinal CMP. Depressive symptoms were associated with weekly spinal CMP but not with weekly chronic local neck pain. Angiotensinogen (AGT) and Fructose-bisphosphate aldolase B (aldolase B) were found to be downregulated in males with weekly spinal CMP compared to healthy controls.

Both Aldolase B and AGT are associated with musculoskeletal issues. AGT is a precursor for various bioactive peptides within the Angiotensin (Ang) system, where Angiotensin-converting enzyme (ACE) plays a key role in converting precursors into active forms of Ang. Initially, AGT is cleaved by renin to form the inactive Ang I, and therefore the complex is often referred to as a renin-angiotensin system (RAS). ACE then converts Ang I into the principal bioactive peptide, Ang II (Balogh *et al*, 2021; Nemoto *et al*, 2023). This conversion is suggested to occur in the circulating blood, possibly explaining why AGT can be identified through blood plasma proteomics.

Recent research has linked the Angiotensin system to pain modulation in inflammatory and neuropathic states. Nemoto et al (2023) found that various Ang peptides appear to affect pain modulation within multiple areas of the CNS. In the brain, Ang receptors are shown to induce hyperalgesia and in the spinal cord, an imbalance of Ang peptides is proposed to cause pain, particularly Ang II is shown to be the key neuromodulator in spinal pain. Within dorsal root ganglia, Ang peptides and their receptors seem to induce hyperalgesia (Nemoto *et al*, 2023). Moreover, the elevated levels of Ang I receptors have been linked to tissue fibrosis in joints and bones, potentially contributing to musculoskeletal problems (Balogh *et al*, 2021; Nemoto *et al*, 2023). Furthermore, circulating Ang II has been shown to promote skeletal muscle atrophy (Powers *et al*, 2018). The signaling of Ang II induces a pathological cascade, resulting in protein degradation and fibrosis in skeletal muscle by elevating the levels of reactive oxygen species (Winslow & Hall, 2019). Ang system-related tissue fibrosis and muscle atrophy might contribute to musculoskeletal problems by weakening the muscles, thereby potentially leading to musculoskeletal pain. Future research may explore the use of AGT as a biomarker for developing targeted drug therapies, since Ang peptide blockers and ACE inhibitors have already been shown to reduce muscle atrophy and sensory neuropathy (Powers *et al*, 2018; Nemoto *et al*, 2023).

Aldolase B is primarily linked to glycolysis and gluconeogenesis. It is expressed mainly in the liver and kidneys (He *et al*, 2020), but Aldolase B can also be detected in blood (The Human Protein Atlas: AldoB, 2024). Recent findings indicate that Aldolase B also impacts the musculoskeletal system: Studies in mice and human carcinoma cells have shown that Aldolase B downregulates the AKT-phosphoinositide-3-kinase (PI3K) pathway (He *et al*, 2020; Gu *et al*, 2023; Peng *et al*, 2023). The AKT/PI3K pathway is crucial for muscle function, as reduced expression may lead to muscle atrophy (Powers *et al*, 2018). Muscle atrophy could contribute to weaker muscles and induce the development of musculoskeletal pain conditions. The association between Aldolase B and CWP has been previously identified in human plasma proteomics research (Olausson *et al*, 2015). In contrast to that study, our research reveals a different perspective on the regulation of Aldolase B, yet the results highlight its importance as a new and meaningful protein marker for CMP. Thus, more studies are needed to understand the association of Aldolase B with CMP as well as CWP.

Local and widespread chronic pain may have different etiology and systemic consequences. Although we were unable to find proteins associated with local neck pain, our study design aimed to compare proteome changes between CMP and localized pain conditions, and not only with healthy controls as has been done previously. Most previous studies focused on middle-aged women, with sample sizes ranging from 15 to 40 participants who experienced pain (Olausson *et al*, 2015; Bäckryd *et al*, 2017; Gerdle *et al*, 2017; Stensson *et al*, 2017; Wåhlén *et al*, 2017, 2018; Gerdle *et al*, 2020, 2022). However, a recent UK Biobank study examined over 20,000 middle-aged individuals (Chen *et al*, 2024). In contrast, our study focused on young adults, whereas in older adults, pain may result from underlying subclinical disease rather than a primary pain syndrome.

Despite having a larger cohort than most comparable studies, we identified only two proteins, both associated with CMP only among males. This may be due to differences in regression methods compared to other studies in the same field and the necessity of generalized linear models due to binomial outcome variables. Our use of regression method enabled accounting for potential confounding factors, which is not possible if less sophisticated approaches are used. As our approach was agnostic and not hypothesis-driven, an extensive number of tests (411) was conducted, nominally significant association may become statistically non-significant when correcting for multiple testing. Thus, it is important to consider the observed effect sizes when comparing our results to others.

Sex, age, depression, and BMI are associated with pain (van Hecke *et al*, 2013). By correcting for all these factors, we improved the possibility of finding relevant associations of proteins with pain. We conclude that considering these four factors strengthens the evidence that the two proteins that we found are robust and relevant and might potentially be truly associated with weekly spinal CMP.

Symptoms of depression often co-occur with chronic pain conditions, and vice versa. In this study, depressive symptoms were linked specifically to multisite pain but not to local neck pain. Earlier studies on depression in CWP show inconsistent findings. One study found similar depression scores between chronic widespread back pain and chronic local back pain groups (Gerhardt *et al*, 2017), while another study found higher depression scores among individuals with chronic pain with a higher percentage of depressive symptoms in widespread pain groups (Bauer *et al*, 2016). Our results suggest that individuals with weekly spinal CMP may have a higher psychological burden than individuals with weekly chronic local neck pain.

Our findings highlight the importance of including both sexes in pain proteomic studies, as pain-associations might be sex-specific, since these associations were found in males but not in females representing the largest group in the regressions. Previous human CWP studies have mainly found protein-pain associations in older menopausal women. They might show great hormonal similarity with men, thus being a potential explanation for why we found results in the young male sample only and not in the young female one. Additionally, sex-based differences in self-reported pain could contribute to these findings. A Swedish study found that women were significantly more likely to report CWP than men (Gerdle *et al*, 2008). Focusing on the frequency rather than the intensity of the pain, may also explain the difference between male and female self-reporting. In a very large study of 1647 individuals Riley et al (2001) found that women with chronic pain are more prone to report higher pain intensity and have pain-related fear and frustration than men with chronic pain (Riley *et al*, 2001). Thus, we speculate that systemic biological effects may manifest easier in men, as they might report pain only when experiencing high-intensity discomfort. Conversely, women may more easily report low-intensity pain, where proteomic changes might be less detectable.

This study also has limitations. Firstly, the definition of CWP used in this study is more restrictive from the one outlined in ICD-11. The focus was primarily on widespread spinal pain, leading to the inclusion of only three pain locations (neck, thorax, and lumbar pain) instead of the standard four. Therefore, we classified the widespread pain in our study as multisite.

Secondly, medication use can potentially influence proteomic measures. The Angiotensin system is commonly known to modulate blood pressure (BP) as the increase in circulating Ang II causes vasoconstriction leading to higher BP (Fyhrquist *et al*, 1995). Thus, theoretically the use of Ang receptor blockers and/or ACE inhibitors could have contributed to the observed decreased AGT levels in males with weekly spinal CMP. However, none of the participants were taking hypertensive medication at the time of this study.

Moreover, proteins with more than 5% of missing values were excluded from the analyses of this study, meaning that only a part of the proteome was scanned, likely comprising the most abundant proteins. Investigating pain-protein associations with low-abundant proteins could lead to the identification of new biomarkers of pain. Yet, such an approach remains challenging in samples like the one that was used in the current study, especially when correcting for multiple testing. Additionally, the removal of high-abundant proteins, which constitute the majority of plasma proteins, may remove low-abundant candidates due to binding interactions, and alternative methods for high-abundant protein removal may offer better sensitivity in identifying low-abundant proteins than the one we used in this study (Palstrøm *et al*, 2020).

Furthermore, the pairwise structure data as twins was not utilized in this study, rather it was corrected as a random intercept in the regressions. In further analyses, the genetic relationship between pain and protein levels could be studied by twin models based on comparing similarities in monozygotic and dizygotic pairs.

Early diagnosis of chronic pain conditions is crucial as the plasticity of the nervous system reduces with increasing age, complicating the pain management. In the future, pain biomarkers could be utilized in the drug discovery and development, clinical trials, pain management and pain diagnosis in comatose, anesthetized, and non-verbal patients (Tracey *et al*, 2019). The observation that proteome level changes are evident only in the wCMP group supports the notion that chronic pain becomes widespread due to more complex biological signaling. Moreover, chronic, and widespread pain conditions are heterogenous diseases making it unlikely to identify a single specific protein associated with the pain condition. Therefore, longitudinal study cohorts are needed in the future to investigate whether changes in protein levels precede CWP or result from it. Additionally, our research findings suggest that including both male and female participants will be highly important in future research.

## Conclusions

In this study, the association of 411 plasma proteins with weekly spinal CMP and weekly local chronic neck pain was investigated using generalized linear mixed-effects models. While no associations were found with the whole sample, interestingly, Aldolase B and Angiotensinogen were downregulated in men with weekly spinal CMP. Both proteins may possibly contribute to the development of musculoskeletal pain and can thus be truly associated with weekly spinal CMP in men. Hence, our findings highlight the importance to include both women and men in the upcoming pain research. Additionally, this study examined depression symptoms in the two pain groups, which revealed that individuals with weekly spinal CMP experienced higher depressive symptoms than the controls.

## Data availability

The data used in the analysis is deposited in the Biobank of the Finnish Institute for Health and Welfare (https://thl.fi/en/web/thl-biobank/forresearchers). It is available to researchers after written application and following the relevant Finnish legislation. To ensure the protection of privacy and compliance with national data protection legislation, a data use/transfer agreement is needed, the content and specific clauses of which will depend on the nature of the requested data.

## Acknowledgements

Mass spectrometry analyses were performed at the Turku Proteomics Facility supported by Biocenter Finland. This project has received funding from the European Union’s Horizon 2020 research and innovation program under grant agreement No 874724. Equal-Life is part of the European Human Exposome Network. Phenotype and omics data collection in FinnTwin12 cohort has been supported by FP7-HEALTH-F4-2007, grant agreement number 201413, National Institute of Alcohol Abuse and Alcoholism (grants AA-12502, AA-00145, and AA-09203 to RJ Rose; AA15416 and K02AA018755 to DM Dick; R01AA015416 to Jessica Salvatore) and the Academy of Finland (grants 100499, 205585, 118555, 141054, 264146, 308248 to JK, and the Centre of Excellence in Complex Disease Genetics (grants 312073, 336823, and 352792 to J Kaprio). J Kaprio acknowledges support from the Academy of Finland (grants 265240, 263278) and the Sigrid Juselius Foundation.

## Author contribution

The study design was developed and discussed by HR, GD, MS, KK and JK. KK and JK provided funding to collect data. KK provided access to proteomic data for analysis. JK provided access to pain data. Processing of FinnTwin12 (pain) and proteomic data was performed by HR. Statistical analyses were performed by HR, GD, and AP. The first version of the manuscript was drafted by HR. All authors participated in the critical revision of the manuscript and approved the final version of the manuscript.

