## Appendix for "Aldolase B and Angiotensinogen are associated with weekly chronic multisite spinal pain in men"

**Supplementary data**

**Appendix 1. The pain questionnaire.** Think about the last three months. How often have you had pain or arching in the following areas?

|  |  | Almost daily | More than once a week | Once a week | Once a month | Rarely  or never |
| --- | --- | --- | --- | --- | --- | --- |
| 1. | Neck or shoulders | 1 | 2 | 3 | 4 | 5 |
| 2. | Upper limbs (arms) |  |  |  |  |  |
| 3. | Chest |  |  |  |  |  |
| 4. | Lower limbs (legs) |  |  |  |  |  |
| 5. | Upper back |  |  |  |  |  |
| 6. | Lower back |  |  |  |  |  |
| 7. | Buttocks |  |  |  |  |  |

**Appendix 2. The shortened (10-item) version of the General Behavior Inventory (GBI).** Is used to measure depressive symptoms.

|  |  | Never | Sometimes | Often | Very often |
| --- | --- | --- | --- | --- | --- |
| 1. | Have you become sad, depressed, or irritable for several days or more without really understanding why? | 1 | 2 | 3 | 4 |
| 2. | Have there been periods of time when you lost almost all interest in the things that you usually like to do (such as hobbies, schoolwork, entertainment)? |  |  |  |  |
| 3. | Have there been periods lasting several days or more when you spent much of your time brooding about unpleasant things that have happened? |  |  |  |  |
| 4. | Have you had periods when you were so down that you found it hard to start talking or that talking took too much energy? |  |  |  |  |
| 5. | Have you experienced several days or more when you were feeling down and depressed, and you also were physically restless, unable to sit still, and had to keep moving or jumping from one activity to another? |  |  |  |  |
| 6. | Have you had periods of sadness and depression when for several days or more, it took you over an hour to get to sleep at night, even though you were very tired? |  |  |  |  |
| 7. | Have there been times of several days or more when you really got down on yourself and felt worthless? |  |  |  |  |
| 8. | Have there been periods of several days or more when you were slowed down and couldn't move as quickly as usual? |  |  |  |  |
| 9. | Have you had long periods when you were down and depressed, interrupted by brief periods when your mood was normal or slightly happy? |  |  |  |  |
| 10. | Have there been periods of time when you felt a persistent sense of gloom? |  |  |  |  |

**Appendix 3. All 411 plasma proteins of the wCMP group in the sensitivity GEE analysis, fully adjusted model.**

| **Protein Accession (ID)** | **Gene** | **Protein** | **Estimate** | **Standard error** | **Nominal p-value** | **Holm adjusted p-value** |
| --- | --- | --- | --- | --- | --- | --- |
| A1L4H1 | SSC5D | Soluble scavenger receptor cysteine-rich domain-containing protein SSC5D | -0.016 | 0.138 | 0.908 | 1 |
| O00187 | MASP2 | Mannan-binding lectin serine protease 2 | 0.126 | 0.142 | 0.374 | 1 |
| O00391 | QSOX1 | Sulfhydryl oxidase 1 | -0.280 | 0.143 | 0.050 | 1 |
| O00533 | CHL1 | Neural cell adhesion molecule L1-like protein | -0.196 | 0.125 | 0.117 | 1 |
| O00592 | PODXL | Podocalyxin | -0.078 | 0.141 | 0.581 | 1 |
| O00602 | FCN1 | Ficolin-1 | -0.174 | 0.143 | 0.223 | 1 |
| O14786 | NRP1 | Neuropilin-1 | -0.112 | 0.138 | 0.415 | 1 |
| O14791 | APOL1 | Apolipoprotein L1 | 0.179 | 0.154 | 0.246 | 1 |
| O15204 | ADAMDEC1 | ADAM DEC1 | -0.005 | 0.138 | 0.972 | 1 |
| O43493 | TGOLN2 | Trans-Golgi network integral membrane protein 2 | -0.152 | 0.127 | 0.230 | 1 |
| O43505 | B4GAT1 | Beta-1,4-glucuronyltransferase 1 | 0.001 | 0.129 | 0.992 | 1 |
| O43852 | CALU | Calumenin | 0.002 | 0.122 | 0.989 | 1 |
| O43866 | CD5L | CD5 antigen-like | -0.178 | 0.140 | 0.203 | 1 |
| O75144 | ICOSLG | ICOS ligand | -0.173 | 0.129 | 0.182 | 1 |
| O75594 | PGLYRP1 | Peptidoglycan recognition protein 1 | -0.203 | 0.125 | 0.103 | 1 |
| O75636 | FCN3 | Ficolin-3 | 0.327 | 0.172 | 0.057 | 1 |
| O75882 | ATRN | Attractin | 0.047 | 0.125 | 0.709 | 1 |
| O94985 | CLSTN1 | Calsyntenin-1 | -0.280 | 0.114 | 0.014 | 1 |
| O95445 | APOM | Apolipoprotein M | 0.106 | 0.132 | 0.423 | 1 |
| O95479 | H6PD | GDH/6PGL endoplasmic bifunctional protein | -0.284 | 0.138 | 0.039 | 1 |
| O95497 | VNN1 | Pantetheinase | 0.193 | 0.149 | 0.195 | 1 |
| P00338 | LDHA | L-lactate dehydrogenase A chain | -0.111 | 0.163 | 0.496 | 1 |
| P00441 | SOD1 | Superoxide dismutase [Cu-Zn] | -0.107 | 0.117 | 0.357 | 1 |
| P00450 | CP | Ceruloplasmin | 0.270 | 0.170 | 0.112 | 1 |
| P00488 | F13A1 | Coagulation factor XIII A chain | 0.057 | 0.126 | 0.653 | 1 |
| P00533 | EGFR | Epidermal growth factor receptor | 0.096 | 0.154 | 0.531 | 1 |
| P00734 | F2 | Prothrombin | -0.211 | 0.130 | 0.103 | 1 |
| P00736 | C1R | Complement C1r subcomponent | -0.119 | 0.128 | 0.353 | 1 |
| P00740 | F9 | Coagulation factor IX | 0.172 | 0.138 | 0.214 | 1 |
| P00742 | F10 | Coagulation factor X | 0.097 | 0.135 | 0.473 | 1 |
| P00746 | CFD | Complement factor D | -0.110 | 0.132 | 0.404 | 1 |
| P00747 | PLG | Plasminogen | 0.087 | 0.151 | 0.566 | 1 |
| P00748 | F12 | Coagulation factor XII | -0.092 | 0.157 | 0.557 | 1 |
| P00751 | CFB | Complement factor B | 0.076 | 0.129 | 0.556 | 1 |
| P00915 | CA1 | Carbonic anhydrase 1 | -0.196 | 0.111 | 0.077 | 1 |
| P00918 | CA2 | Carbonic anhydrase 2 | 0.080 | 0.131 | 0.542 | 1 |
| P01008 | SERPINC1 | Antithrombin-III | -0.064 | 0.143 | 0.653 | 1 |
| P01011 | SERPINA3 | Alpha-1-antichymotrypsin | -0.239 | 0.133 | 0.071 | 1 |
| P01019 | AGT | Angiotensinogen | 0.125 | 0.156 | 0.422 | 1 |
| P01024 | C3 | Complement C3 | -0.084 | 0.132 | 0.525 | 1 |
| P01031 | C5 | Complement C5 | 0.085 | 0.130 | 0.515 | 1 |
| P01033 | TIMP1 | Metalloproteinase inhibitor 1 | 0.002 | 0.131 | 0.989 | 1 |
| P01034 | CST3 | Cystatin-C | 0.139 | 0.136 | 0.307 | 1 |
| P01042 | KNG1 | Kininogen-1 | 0.029 | 0.153 | 0.847 | 1 |
| P01137 | TGFB1 | Transforming growth factor beta-1 proprotein | -0.031 | 0.135 | 0.821 | 1 |
| P01344 | IGF2 | Insulin-like growth factor II | 0.181 | 0.125 | 0.149 | 1 |
| P02042 | HBD | Hemoglobin subunit delta | 0.031 | 0.119 | 0.791 | 1 |
| P02144 | MB | Myoglobin | -0.135 | 0.130 | 0.300 | 1 |
| P02452 | COL1A1 | Collagen alpha-1(I) chain | 0.035 | 0.133 | 0.793 | 1 |
| P02533 | KRT14 | Keratin, type I cytoskeletal 14 | 0.132 | 0.165 | 0.423 | 1 |
| P02649 | APOE | Apolipoprotein E | -0.024 | 0.144 | 0.866 | 1 |
| P02652 | APOA2 | Apolipoprotein A-II | 0.004 | 0.145 | 0.977 | 1 |
| P02654 | APOC1 | Apolipoprotein C-I | 0.072 | 0.127 | 0.571 | 1 |
| P02655 | APOC2 | Apolipoprotein C-II | -0.026 | 0.134 | 0.844 | 1 |
| P02656 | APOC3 | Apolipoprotein C-III | 0.082 | 0.128 | 0.522 | 1 |
| P02741 | CRP | C-reactive protein | -0.024 | 0.144 | 0.866 | 1 |
| P02743 | APCS | Serum amyloid P-component | 0.121 | 0.144 | 0.401 | 1 |
| P02745 | C1QA | Complement C1q subcomponent subunit A | 0.062 | 0.130 | 0.632 | 1 |
| P02746 | C1QB | Complement C1q subcomponent subunit B | 0.030 | 0.128 | 0.815 | 1 |
| P02747 | C1QC | Complement C1q subcomponent subunit C | 0.041 | 0.120 | 0.732 | 1 |
| P02748 | C9 | Complement component C9 | -0.173 | 0.128 | 0.177 | 1 |
| P02749 | APOH | Beta-2-glycoprotein 1 | 0.139 | 0.145 | 0.337 | 1 |
| P02750 | LRG1 | Leucine-rich alpha-2-glycoprotein | -0.133 | 0.158 | 0.397 | 1 |
| P02751 | FN1 | Fibronectin | -0.013 | 0.143 | 0.925 | 1 |
| P02753 | RBP4 | Retinol-binding protein 4 | 0.190 | 0.128 | 0.138 | 1 |
| P02760 | AMBP | Protein AMBP | -0.088 | 0.140 | 0.531 | 1 |
| P02765 | AHSG | Alpha-2-HS-glycoprotein | 0.205 | 0.144 | 0.153 | 1 |
| P02766 | TTR | Transthyretin | 0.083 | 0.141 | 0.555 | 1 |
| P02774 | GC | Vitamin D-binding protein | 0.233 | 0.134 | 0.084 | 1 |
| P02775 | PPBP | Platelet basic protein | -0.186 | 0.132 | 0.158 | 1 |
| P02776 | PF4 | Platelet factor 4 | -0.098 | 0.129 | 0.450 | 1 |
| P02790 | HPX | Hemopexin | -0.015 | 0.127 | 0.904 | 1 |
| P03950 | ANG | Angiogenin | -0.051 | 0.121 | 0.675 | 1 |
| P03951 | F11 | Coagulation factor XI | -0.061 | 0.130 | 0.638 | 1 |
| P03952 | KLKB1 | Plasma kallikrein | -0.051 | 0.130 | 0.695 | 1 |
| P03973 | SLPI | Antileukoproteinase | -0.096 | 0.132 | 0.467 | 1 |
| P04003 | C4BPA | C4b-binding protein alpha chain | -0.243 | 0.141 | 0.084 | 1 |
| P04004 | VTN | Vitronectin | 0.150 | 0.147 | 0.305 | 1 |
| P04040 | CAT | Catalase | -0.042 | 0.127 | 0.741 | 1 |
| P04070 | PROC | Vitamin K-dependent protein C | 0.232 | 0.142 | 0.102 | 1 |
| P04075 | ALDOA | Fructose-bisphosphate aldolase A | -0.162 | 0.145 | 0.263 | 1 |
| P04114 | APOB | Apolipoprotein B-100 | 0.044 | 0.138 | 0.749 | 1 |
| P04180 | LCAT | Phosphatidylcholine-sterol acyltransferase | 0.139 | 0.142 | 0.328 | 1 |
| P04196 | HRG | Histidine-rich glycoprotein | -0.156 | 0.134 | 0.244 | 1 |
| P04217 | A1BG | Alpha-1B-glycoprotein | -0.065 | 0.147 | 0.657 | 1 |
| P04259 | KRT6B | Keratin, type II cytoskeletal 6B | -0.097 | 0.128 | 0.448 | 1 |
| P04264 | KRT1 | Keratin, type II cytoskeletal 1 | 0.052 | 0.138 | 0.704 | 1 |
| P04275 | VWF | von Willebrand factor | -0.148 | 0.132 | 0.262 | 1 |
| P04278 | SHBG | Sex hormone-binding globulin | 0.118 | 0.196 | 0.547 | 1 |
| P04406 | GAPDH | Glyceraldehyde-3-phosphate dehydrogenase | 0.092 | 0.130 | 0.481 | 1 |
| P04439 | HLA-A | HLA class I histocompatibility antigen, A alpha chain | -0.134 | 0.134 | 0.317 | 1 |
| P04746 | AMY2A | Pancreatic alpha-amylase | -0.073 | 0.139 | 0.600 | 1 |
| P05019 | IGF1 | Insulin-like growth factor I | -0.156 | 0.137 | 0.255 | 1 |
| P05062 | ALDOB | Fructose-bisphosphate aldolase B | -0.260 | 0.135 | 0.054 | 1 |
| P05067 | APP | Amyloid-beta precursor protein | -0.005 | 0.143 | 0.973 | 1 |
| P05090 | APOD | Apolipoprotein D | -0.083 | 0.142 | 0.559 | 1 |
| P05106 | ITGB3 | Integrin beta-3 | 0.042 | 0.130 | 0.746 | 1 |
| P05109 | S100A8 | Protein S100-A8 | 0.198 | 0.157 | 0.208 | 1 |
| P05121 | SERPINE1 | Plasminogen activator inhibitor 1 | -0.052 | 0.124 | 0.672 | 1 |
| P05154 | SERPINA5 | Plasma serine protease inhibitor | -0.252 | 0.131 | 0.053 | 1 |
| P05155 | SERPING1 | Plasma protease C1 inhibitor | -0.078 | 0.128 | 0.542 | 1 |
| P05156 | CFI | Complement factor I | 0.061 | 0.143 | 0.671 | 1 |
| P05160 | F13B | Coagulation factor XIII B chain | 0.120 | 0.143 | 0.400 | 1 |
| P05164 | MPO | Myeloperoxidase | 0.049 | 0.135 | 0.716 | 1 |
| P05362 | ICAM1 | Intercellular adhesion molecule 1 | -0.026 | 0.123 | 0.830 | 1 |
| P05451 | REG1A | Lithostathine-1-alpha | -0.113 | 0.126 | 0.369 | 1 |
| P05452 | CLEC3B | Tetranectin | -0.017 | 0.142 | 0.908 | 1 |
| P05543 | SERPINA7 | Thyroxine-binding globulin | 0.289 | 0.155 | 0.062 | 1 |
| P05546 | SERPIND1 | Heparin cofactor 2 | 0.186 | 0.140 | 0.183 | 1 |
| P05556 | ITGB1 | Integrin beta-1 | 0.140 | 0.141 | 0.319 | 1 |
| P06276 | BCHE | Cholinesterase | -0.076 | 0.158 | 0.629 | 1 |
| P06396 | GSN | Gelsolin | 0.024 | 0.148 | 0.870 | 1 |
| P06681 | C2 | Complement C2 | -0.007 | 0.129 | 0.958 | 1 |
| P06702 | S100A9 | Protein S100-A9 | 0.054 | 0.153 | 0.722 | 1 |
| P06727 | APOA4 | Apolipoprotein A-IV | 0.073 | 0.147 | 0.620 | 1 |
| P06732 | CKM | Creatine kinase M-type | 0.104 | 0.155 | 0.503 | 1 |
| P07195 | LDHB | L-lactate dehydrogenase B chain | -0.107 | 0.133 | 0.421 | 1 |
| P07225 | PROS1 | Vitamin K-dependent protein S | -0.025 | 0.151 | 0.869 | 1 |
| P07237 | P4HB | Protein disulfide-isomerase | 0.099 | 0.195 | 0.613 | 1 |
| P07333 | CSF1R | Macrophage colony-stimulating factor 1 receptor | -0.163 | 0.129 | 0.206 | 1 |
| P07339 | CTSD | Cathepsin D | -0.084 | 0.138 | 0.541 | 1 |
| P07357 | C8A | Complement component C8 alpha chain | 0.064 | 0.128 | 0.617 | 1 |
| P07358 | C8B | Complement component C8 beta chain | 0.090 | 0.130 | 0.492 | 1 |
| P07359 | GP1BA | Platelet glycoprotein Ib alpha chain | -0.158 | 0.130 | 0.225 | 1 |
| P07360 | C8G | Complement component C8 gamma chain | 0.161 | 0.140 | 0.252 | 1 |
| P07437 | TUBB | Tubulin beta chain | 0.162 | 0.142 | 0.253 | 1 |
| P07602 | PSAP | Prosaposin | 0.152 | 0.144 | 0.291 | 1 |
| P07737 | PFN1 | Profilin-1 | -0.087 | 0.150 | 0.562 | 1 |
| P07911 | UMOD | Uromodulin | -0.033 | 0.118 | 0.777 | 1 |
| P07942 | LAMB1 | Laminin subunit beta-1 | -0.222 | 0.132 | 0.093 | 1 |
| P07951 | TPM2 | Tropomyosin beta chain | -0.023 | 0.118 | 0.843 | 1 |
| P07996 | THBS1 | Thrombospondin-1 | 0.052 | 0.123 | 0.673 | 1 |
| P07998 | RNASE1 | Ribonuclease pancreatic | -0.158 | 0.121 | 0.193 | 1 |
| P08185 | SERPINA6 | Corticosteroid-binding globulin | 0.127 | 0.165 | 0.440 | 1 |
| P08195 | SLC3A2 | 4F2 cell-surface antigen heavy chain | -0.237 | 0.149 | 0.112 | 1 |
| P08238 | HSP90AB1 | Heat shock protein HSP 90-beta | -0.157 | 0.113 | 0.165 | 1 |
| P08253 | MMP2 | 72 kDa type IV collagenase | -0.307 | 0.124 | 0.013 | 1 |
| P08294 | SOD3 | Extracellular superoxide dismutase [Cu-Zn] | -0.055 | 0.127 | 0.662 | 1 |
| P08311 | CTSG | Cathepsin G | -0.151 | 0.116 | 0.192 | 1 |
| P08493 | MGP | Matrix Gla protein | -0.022 | 0.123 | 0.860 | 1 |
| P08514 | ITGA2B | Integrin alpha-IIb | -0.473 | 0.134 | 0.000 | 0.1630851 |
| P08519 | LPA | Apolipoprotein(a) | 0.099 | 0.117 | 0.397 | 1 |
| P08571 | CD14 | Monocyte differentiation antigen CD14 | 0.132 | 0.151 | 0.382 | 1 |
| P08603 | CFH | Complement factor H | -0.111 | 0.131 | 0.395 | 1 |
| P08697 | SERPINF2 | Alpha-2-antiplasmin | 0.108 | 0.128 | 0.399 | 1 |
| P08709 | F7 | Coagulation factor VII | 0.169 | 0.136 | 0.213 | 1 |
| P08779 | KRT16 | Keratin, type I cytoskeletal 16 | 0.191 | 0.130 | 0.141 | 1 |
| P08887 | IL6R | Interleukin-6 receptor subunit alpha | 0.023 | 0.143 | 0.872 | 1 |
| P09172 | DBH | Dopamine beta-hydroxylase | 0.037 | 0.126 | 0.768 | 1 |
| P09486 | SPARC | SPARC | -0.063 | 0.154 | 0.682 | 1 |
| P09871 | C1S | Complement C1s subcomponent | 0.171 | 0.123 | 0.163 | 1 |
| P09960 | LTA4H | Leukotriene A-4 hydrolase | 0.084 | 0.129 | 0.515 | 1 |
| P0C0L4 | C4A | Complement C4-A | 0.014 | 0.138 | 0.922 | 1 |
| P0C0L5 | C4B | Complement C4-B | 0.363 | 0.145 | 0.012 | 1 |
| P0DJI8 | SAA1 | Serum amyloid A-1 protein | -0.172 | 0.134 | 0.202 | 1 |
| P10124 | SRGN | Serglycin | -0.103 | 0.132 | 0.435 | 1 |
| P10153 | RNASE2 | Non-secretory ribonuclease | 0.079 | 0.135 | 0.561 | 1 |
| P10586 | PTPRF | Receptor-type tyrosine-protein phosphatase F | 0.161 | 0.151 | 0.286 | 1 |
| P10643 | C7 | Complement component C7 | -0.007 | 0.129 | 0.954 | 1 |
| P10646 | TFPI | Tissue factor pathway inhibitor | 0.080 | 0.123 | 0.515 | 1 |
| P10720 | PF4V1 | Platelet factor 4 variant | 0.050 | 0.126 | 0.692 | 1 |
| P10721 | KIT | Mast/stem cell growth factor receptor Kit | 0.225 | 0.126 | 0.076 | 1 |
| P10909 | CLU | Clusterin | 0.036 | 0.130 | 0.781 | 1 |
| P11021 | HSPA5 | Endoplasmic reticulum chaperone BiP | -0.091 | 0.142 | 0.519 | 1 |
| P11142 | HSPA8 | Heat shock cognate 71 kDa protein | -0.102 | 0.130 | 0.430 | 1 |
| P11226 | MBL2 | Mannose-binding protein C | -0.294 | 0.122 | 0.016 | 1 |
| P11279 | LAMP1 | Lysosome-associated membrane glycoprotein 1 | 0.084 | 0.129 | 0.514 | 1 |
| P11362 | FGFR1 | Fibroblast growth factor receptor 1 | -0.134 | 0.122 | 0.271 | 1 |
| P11597 | CETP | Cholesteryl ester transfer protein | 0.099 | 0.138 | 0.474 | 1 |
| P11717 | IGF2R | Cation-independent mannose-6-phosphate receptor | -0.117 | 0.134 | 0.380 | 1 |
| P12109 | COL6A1 | Collagen alpha-1(VI) chain | -0.341 | 0.136 | 0.012 | 1 |
| P12111 | COL6A3 | Collagen alpha-3(VI) chain | -0.225 | 0.125 | 0.071 | 1 |
| P12259 | F5 | Coagulation factor V | 0.007 | 0.137 | 0.962 | 1 |
| P12821 | ACE | Angiotensin-converting enzyme | -0.072 | 0.129 | 0.577 | 1 |
| P12830 | CDH1 | Cadherin-1 | -0.192 | 0.134 | 0.154 | 1 |
| P12955 | PEPD | Xaa-Pro dipeptidase | -0.088 | 0.136 | 0.520 | 1 |
| P13473 | LAMP2 | Lysosome-associated membrane glycoprotein 2 | 0.037 | 0.138 | 0.792 | 1 |
| P13591 | NCAM1 | Neural cell adhesion molecule 1 | -0.014 | 0.121 | 0.911 | 1 |
| P13598 | ICAM2 | Intercellular adhesion molecule 2 | -0.356 | 0.132 | 0.007 | 1 |
| P13645 | KRT10 | Keratin, type I cytoskeletal 10 | -0.041 | 0.141 | 0.772 | 1 |
| P13646 | KRT13 | Keratin, type I cytoskeletal 13 | -0.163 | 0.128 | 0.204 | 1 |
| P13671 | C6 | Complement component C6 | -0.052 | 0.137 | 0.704 | 1 |
| P13727 | PRG2 | Bone marrow proteoglycan | -0.117 | 0.131 | 0.374 | 1 |
| P13796 | LCP1 | Plastin-2 | 0.061 | 0.145 | 0.673 | 1 |
| P14151 | SELL | L-selectin | 0.012 | 0.128 | 0.922 | 1 |
| P14209 | CD99 | CD99 antigen | -0.112 | 0.136 | 0.407 | 1 |
| P14314 | PRKCSH | Glucosidase 2 subunit beta | -0.041 | 0.126 | 0.745 | 1 |
| P14543 | NID1 | Nidogen-1 | -0.094 | 0.125 | 0.449 | 1 |
| P14618 | PKM | Pyruvate kinase PKM | -0.202 | 0.144 | 0.160 | 1 |
| P14625 | HSP90B1 | Endoplasmin | -0.167 | 0.126 | 0.186 | 1 |
| P14780 | MMP9 | Matrix metalloproteinase-9 | -0.112 | 0.141 | 0.430 | 1 |
| P15144 | ANPEP | Aminopeptidase N | -0.162 | 0.131 | 0.216 | 1 |
| P15151 | PVR | Poliovirus receptor | 0.099 | 0.139 | 0.475 | 1 |
| P15169 | CPN1 | Carboxypeptidase N catalytic chain | 0.062 | 0.137 | 0.654 | 1 |
| P16035 | TIMP2 | Metalloproteinase inhibitor 2 | -0.194 | 0.125 | 0.121 | 1 |
| P16070 | CD44 | CD44 antigen | -0.245 | 0.126 | 0.053 | 1 |
| P16112 | ACAN | Aggrecan core protein | -0.106 | 0.136 | 0.434 | 1 |
| P16930 | FAH | Fumarylacetoacetase | -0.047 | 0.128 | 0.713 | 1 |
| P17813 | ENG | Endoglin | -0.165 | 0.136 | 0.225 | 1 |
| P17936 | IGFBP3 | Insulin-like growth factor-binding protein 3 | 0.078 | 0.133 | 0.555 | 1 |
| P18065 | IGFBP2 | Insulin-like growth factor-binding protein 2 | -0.124 | 0.133 | 0.350 | 1 |
| P18206 | VCL | Vinculin | 0.033 | 0.132 | 0.805 | 1 |
| P18428 | LBP | Lipopolysaccharide-binding protein | 0.122 | 0.131 | 0.350 | 1 |
| P19021 | PAM | Peptidyl-glycine alpha-amidating monooxygenase | 0.064 | 0.136 | 0.641 | 1 |
| P19022 | CDH2 | Cadherin-2 | 0.177 | 0.132 | 0.179 | 1 |
| P19320 | VCAM1 | Vascular cell adhesion protein 1 | -0.137 | 0.132 | 0.298 | 1 |
| P19823 | ITIH2 | Inter-alpha-trypsin inhibitor heavy chain H2 | -0.006 | 0.148 | 0.965 | 1 |
| P19827 | ITIH1 | Inter-alpha-trypsin inhibitor heavy chain H1 | 0.082 | 0.150 | 0.585 | 1 |
| P20023 | CR2 | Complement receptor type 2 | 0.079 | 0.135 | 0.561 | 1 |
| P20742 | PZP | Pregnancy zone protein | -0.022 | 0.166 | 0.894 | 1 |
| P20851 | C4BPB | C4b-binding protein beta chain | -0.210 | 0.142 | 0.139 | 1 |
| P21333 | FLNA | Filamin-A | -0.157 | 0.141 | 0.265 | 1 |
| P22105 | TNXB | Tenascin-X | -0.243 | 0.133 | 0.068 | 1 |
| P22352 | GPX3 | Glutathione peroxidase 3 | 0.098 | 0.134 | 0.467 | 1 |
| P22692 | IGFBP4 | Insulin-like growth factor-binding protein 4 | 0.227 | 0.154 | 0.141 | 1 |
| P22792 | CPN2 | Carboxypeptidase N subunit 2 | 0.109 | 0.134 | 0.417 | 1 |
| P22891 | PROZ | Vitamin K-dependent protein Z | 0.082 | 0.144 | 0.572 | 1 |
| P22897 | MRC1 | Macrophage mannose receptor 1 | 0.147 | 0.122 | 0.230 | 1 |
| P23142 | FBLN1 | Fibulin-1 | 0.030 | 0.123 | 0.807 | 1 |
| P23470 | PTPRG | Receptor-type tyrosine-protein phosphatase gamma | -0.179 | 0.136 | 0.190 | 1 |
| P23528 | CFL1 | Cofilin-1 | 0.025 | 0.132 | 0.849 | 1 |
| P24043 | LAMA2 | Laminin subunit alpha-2 | -0.078 | 0.124 | 0.528 | 1 |
| P24592 | IGFBP6 | Insulin-like growth factor-binding protein 6 | 0.016 | 0.129 | 0.902 | 1 |
| P24593 | IGFBP5 | Insulin-like growth factor-binding protein 5 | 0.086 | 0.124 | 0.491 | 1 |
| P24821 | TNC | Tenascin | -0.111 | 0.126 | 0.379 | 1 |
| P25311 | AZGP1 | Zinc-alpha-2-glycoprotein | 0.037 | 0.146 | 0.799 | 1 |
| P25774 | CTSS | Cathepsin S | -0.158 | 0.147 | 0.281 | 1 |
| P26038 | MSN | Moesin | -0.016 | 0.137 | 0.906 | 1 |
| P26927 | MST1 | Hepatocyte growth factor-like protein | 0.113 | 0.123 | 0.357 | 1 |
| P27169 | PON1 | Serum paraoxonase/arylesterase 1 | -0.006 | 0.126 | 0.962 | 1 |
| P27487 | DPP4 | Dipeptidyl peptidase 4 | -0.033 | 0.133 | 0.805 | 1 |
| P27797 | CALR | Calreticulin | -0.102 | 0.132 | 0.437 | 1 |
| P27918 | CFP | Properdin | 0.155 | 0.136 | 0.252 | 1 |
| P28799 | GRN | Progranulin | 0.111 | 0.137 | 0.420 | 1 |
| P29401 | TKT | Transketolase | -0.195 | 0.152 | 0.198 | 1 |
| P29622 | SERPINA4 | Kallistatin | -0.017 | 0.142 | 0.907 | 1 |
| P30043 | BLVRB | Flavin reductase (NADPH) | -0.112 | 0.131 | 0.393 | 1 |
| P30101 | PDIA3 | Protein disulfide-isomerase A3 | 0.059 | 0.120 | 0.625 | 1 |
| P32119 | PRDX2 | Peroxiredoxin-2 | -0.045 | 0.123 | 0.714 | 1 |
| P32942 | ICAM3 | Intercellular adhesion molecule 3 | -0.092 | 0.134 | 0.493 | 1 |
| P33151 | CDH5 | Cadherin-5 | -0.230 | 0.138 | 0.095 | 1 |
| P33908 | MAN1A1 | Mannosyl-oligosaccharide 1,2-alpha-mannosidase IA | 0.069 | 0.133 | 0.605 | 1 |
| P34096 | RNASE4 | Ribonuclease 4 | -0.119 | 0.134 | 0.377 | 1 |
| P35030 | PRSS3 | Trypsin-3 | -0.004 | 0.126 | 0.973 | 1 |
| P35443 | THBS4 | Thrombospondin-4 | -0.131 | 0.138 | 0.339 | 1 |
| P35527 | KRT9 | Keratin, type I cytoskeletal 9 | -0.012 | 0.127 | 0.922 | 1 |
| P35542 | SAA4 | Serum amyloid A-4 protein | -0.317 | 0.130 | 0.015 | 1 |
| P35555 | FBN1 | Fibrillin-1 | 0.163 | 0.138 | 0.238 | 1 |
| P35858 | IGFALS | Insulin-like growth factor-binding protein complex acid labile subunit | 0.232 | 0.133 | 0.080 | 1 |
| P35908 | KRT2 | Keratin, type II cytoskeletal 2 epidermal | 0.181 | 0.139 | 0.191 | 1 |
| P35916 | FLT4 | Vascular endothelial growth factor receptor 3 | -0.015 | 0.127 | 0.903 | 1 |
| P36955 | SERPINF1 | Pigment epithelium-derived factor | 0.363 | 0.150 | 0.015 | 1 |
| P36980 | CFHR2 | Complement factor H-related protein 2 | 0.351 | 0.176 | 0.046 | 1 |
| P39060 | COL18A1 | Collagen alpha-1(XVIII) chain | 0.032 | 0.128 | 0.805 | 1 |
| P40189 | IL6ST | Interleukin-6 receptor subunit beta | -0.129 | 0.138 | 0.348 | 1 |
| P40197 | GP5 | Platelet glycoprotein V | 0.036 | 0.124 | 0.773 | 1 |
| P41222 | PTGDS | Prostaglandin-H2 D-isomerase | -0.084 | 0.129 | 0.512 | 1 |
| P43121 | MCAM | Cell surface glycoprotein MUC18 | 0.041 | 0.125 | 0.742 | 1 |
| P43251 | BTD | Biotinidase | -0.052 | 0.140 | 0.709 | 1 |
| P43652 | AFM | Afamin | -0.027 | 0.128 | 0.835 | 1 |
| P48594 | SERPINB4 | Serpin B4 | -0.084 | 0.135 | 0.534 | 1 |
| P48668 | KRT6C | Keratin, type II cytoskeletal 6C | 0.061 | 0.132 | 0.644 | 1 |
| P48740 | MASP1 | Mannan-binding lectin serine protease 1 | 0.029 | 0.139 | 0.834 | 1 |
| P49747 | COMP | Cartilage oligomeric matrix protein | -0.009 | 0.136 | 0.946 | 1 |
| P49908 | SELENOP | Selenoprotein P | -0.009 | 0.145 | 0.949 | 1 |
| P49913 | CAMP | Cathelicidin antimicrobial peptide | -0.054 | 0.132 | 0.683 | 1 |
| P50395 | GDI2 | Rab GDP dissociation inhibitor beta | 0.020 | 0.135 | 0.882 | 1 |
| P51884 | LUM | Lumican | 0.066 | 0.131 | 0.615 | 1 |
| P53634 | CTSC | Dipeptidyl peptidase 1 | -0.181 | 0.136 | 0.182 | 1 |
| P54289 | CACNA2D1 | Voltage-dependent calcium channel subunit alpha-2/delta-1 | -0.067 | 0.142 | 0.637 | 1 |
| P54802 | NAGLU | Alpha-N-acetylglucosaminidase | 0.063 | 0.135 | 0.643 | 1 |
| P55056 | APOC4 | Apolipoprotein C-IV | 0.225 | 0.149 | 0.131 | 1 |
| P55058 | PLTP | Phospholipid transfer protein | -0.078 | 0.132 | 0.557 | 1 |
| P55103 | INHBC | Inhibin beta C chain | 0.125 | 0.132 | 0.343 | 1 |
| P55290 | CDH13 | Cadherin-13 | -0.098 | 0.123 | 0.426 | 1 |
| P59666 | DEFA3 | Neutrophil defensin 3 | -0.223 | 0.155 | 0.150 | 1 |
| P60174 | TPI1 | Triosephosphate isomerase | -0.187 | 0.179 | 0.296 | 1 |
| P60709 | ACTB | Actin, cytoplasmic 1 | 0.057 | 0.134 | 0.672 | 1 |
| P61224 | RAP1B | Ras-related protein Rap-1b | 0.022 | 0.120 | 0.858 | 1 |
| P61626 | LYZ | Lysozyme C | 0.100 | 0.135 | 0.460 | 1 |
| P61769 | B2M | Beta-2-microglobulin | -0.132 | 0.130 | 0.311 | 1 |
| P62328 | TMSB4X | Thymosin beta-4 | -0.037 | 0.125 | 0.764 | 1 |
| P62937 | PPIA | Peptidyl-prolyl cis-trans isomerase A | -0.035 | 0.126 | 0.779 | 1 |
| P63104 | YWHAZ | 14-3-3 protein zeta/delta | 0.065 | 0.149 | 0.662 | 1 |
| P63261 | ACTG1 | Actin, cytoplasmic 2 | -0.099 | 0.137 | 0.470 | 1 |
| P68032.P68133 | ACTC1;ACTA1 | Actin, alpha cardiac muscle 1;Actin, alpha skeletal muscle | 0.098 | 0.127 | 0.440 | 1 |
| P68363 | TUBA1B | Tubulin alpha-1B chain | 0.209 | 0.141 | 0.137 | 1 |
| P68871 | HBB | Hemoglobin subunit beta | -0.072 | 0.118 | 0.542 | 1 |
| P69892 | HBG2 | Hemoglobin subunit gamma-2 | -0.047 | 0.129 | 0.716 | 1 |
| P69905 | HBA1 | Hemoglobin subunit alpha | -0.066 | 0.120 | 0.582 | 1 |
| P78509 | RELN | Reelin | -0.253 | 0.130 | 0.051 | 1 |
| P80108 | GPLD1 | Phosphatidylinositol-glycan-specific phospholipase D | 0.026 | 0.147 | 0.857 | 1 |
| P80723 | BASP1 | Brain acid soluble protein 1 | 0.056 | 0.120 | 0.642 | 1 |
| P98160 | HSPG2 | Basement membrane-specific heparan sulfate proteoglycan core protein | -0.060 | 0.139 | 0.664 | 1 |
| Q00610 | CLTC | Clathrin heavy chain 1 | 0.183 | 0.154 | 0.232 | 1 |
| Q01459 | CTBS | Di-N-acetylchitobiase | -0.028 | 0.135 | 0.834 | 1 |
| Q02985 | CFHR3 | Complement factor H-related protein 3 | 0.365 | 0.147 | 0.013 | 1 |
| Q03591 | CFHR1 | Complement factor H-related protein 1 | 0.126 | 0.146 | 0.385 | 1 |
| Q04721 | NOTCH2 | Neurogenic locus notch homolog protein 2 | 0.014 | 0.139 | 0.918 | 1 |
| Q04756 | HGFAC | Hepatocyte growth factor activator | -0.075 | 0.136 | 0.581 | 1 |
| Q06033 | ITIH3 | Inter-alpha-trypsin inhibitor heavy chain H3 | -0.364 | 0.141 | 0.010 | 1 |
| Q06830 | PRDX1 | Peroxiredoxin-1 | 0.073 | 0.138 | 0.596 | 1 |
| Q07954 | LRP1 | Prolow-density lipoprotein receptor-related protein 1 | 0.038 | 0.125 | 0.760 | 1 |
| Q08380 | LGALS3BP | Galectin-3-binding protein | 0.230 | 0.146 | 0.116 | 1 |
| Q10588 | BST1 | ADP-ribosyl cyclase/cyclic ADP-ribose hydrolase 2 | -0.018 | 0.130 | 0.893 | 1 |
| Q12805 | EFEMP1 | EGF-containing fibulin-like extracellular matrix protein 1 | 0.170 | 0.129 | 0.190 | 1 |
| Q12841 | FSTL1 | Follistatin-related protein 1 | 0.004 | 0.117 | 0.973 | 1 |
| Q12860 | CNTN1 | Contactin-1 | -0.253 | 0.128 | 0.048 | 1 |
| Q12884 | FAP | Prolyl endopeptidase FAP | -0.038 | 0.133 | 0.776 | 1 |
| Q12907 | LMAN2 | Vesicular integral-membrane protein VIP36 | -0.155 | 0.137 | 0.259 | 1 |
| Q12913 | PTPRJ | Receptor-type tyrosine-protein phosphatase eta | 0.204 | 0.140 | 0.146 | 1 |
| Q13093 | PLA2G7 | Platelet-activating factor acetylhydrolase | 0.036 | 0.136 | 0.789 | 1 |
| Q13103 | SPP2 | Secreted phosphoprotein 24 | -0.110 | 0.139 | 0.427 | 1 |
| Q13201 | MMRN1 | Multimerin-1 | -0.125 | 0.113 | 0.272 | 1 |
| Q13332 | PTPRS | Receptor-type tyrosine-protein phosphatase S | -0.073 | 0.128 | 0.571 | 1 |
| Q13449 | LSAMP | Limbic system-associated membrane protein | -0.400 | 0.143 | 0.005 | 1 |
| Q13740 | ALCAM | CD166 antigen | -0.374 | 0.142 | 0.008 | 1 |
| Q13790 | APOF | Apolipoprotein F | 0.141 | 0.138 | 0.306 | 1 |
| Q13822 | ENPP2 | Ectonucleotide pyrophosphatase/phosphodiesterase family member 2 | 0.189 | 0.152 | 0.213 | 1 |
| Q14112 | NID2 | Nidogen-2 | -0.071 | 0.135 | 0.600 | 1 |
| Q14126 | DSG2 | Desmoglein-2 | -0.004 | 0.125 | 0.978 | 1 |
| Q14314 | FGL2 | Fibroleukin | -0.156 | 0.114 | 0.171 | 1 |
| Q14515 | SPARCL1 | SPARC-like protein 1 | 0.065 | 0.133 | 0.627 | 1 |
| Q14520 | HABP2 | Hyaluronan-binding protein 2 | -0.039 | 0.140 | 0.779 | 1 |
| Q14624 | ITIH4 | Inter-alpha-trypsin inhibitor heavy chain H4 | 0.013 | 0.155 | 0.935 | 1 |
| Q14766 | LTBP1 | Latent-transforming growth factor beta-binding protein 1 | -0.039 | 0.125 | 0.755 | 1 |
| Q14956 | GPNMB | Transmembrane glycoprotein NMB | -0.140 | 0.130 | 0.282 | 1 |
| Q15063 | POSTN | Periostin | -0.168 | 0.126 | 0.184 | 1 |
| Q15113 | PCOLCE | Procollagen C-endopeptidase enhancer 1 | -0.108 | 0.139 | 0.440 | 1 |
| Q15166 | PON3 | Serum paraoxonase/lactonase 3 | 0.116 | 0.142 | 0.414 | 1 |
| Q15485 | FCN2 | Ficolin-2 | 0.034 | 0.130 | 0.794 | 1 |
| Q15582 | TGFBI | Transforming growth factor-beta-induced protein ig-h3 | 0.068 | 0.139 | 0.624 | 1 |
| Q15828 | CST6 | Cystatin-M | -0.105 | 0.122 | 0.389 | 1 |
| Q15848 | ADIPOQ | Adiponectin | 0.037 | 0.143 | 0.798 | 1 |
| Q16270 | IGFBP7 | Insulin-like growth factor-binding protein 7 | -0.282 | 0.127 | 0.027 | 1 |
| Q16610 | ECM1 | Extracellular matrix protein 1 | 0.003 | 0.133 | 0.981 | 1 |
| Q16706 | MAN2A1 | Alpha-mannosidase 2 | -0.009 | 0.119 | 0.939 | 1 |
| Q16853 | AOC3 | Membrane primary amine oxidase | -0.158 | 0.127 | 0.214 | 1 |
| Q4LDE5 | SVEP1 | Sushi, von Willebrand factor type A, EGF and pentraxin domain-containing protein 1 | -0.221 | 0.126 | 0.080 | 1 |
| Q562R1 | ACTBL2 | Beta-actin-like protein 2 | -0.142 | 0.113 | 0.209 | 1 |
| Q6EMK4 | VASN | Vasorin | -0.211 | 0.141 | 0.135 | 1 |
| Q6UWP8 | SBSN | Suprabasin | -0.134 | 0.127 | 0.289 | 1 |
| Q6UX71 | PLXDC2 | Plexin domain-containing protein 2 | -0.141 | 0.141 | 0.318 | 1 |
| Q6UXB8 | PI16 | Peptidase inhibitor 16 | -0.212 | 0.127 | 0.095 | 1 |
| Q6UY14 | ADAMTSL4 | ADAMTS-like protein 4 | -0.108 | 0.119 | 0.361 | 1 |
| Q6YHK3 | CD109 | CD109 antigen | 0.000 | 0.131 | 0.997 | 1 |
| Q76LX8 | ADAMTS13 | A disintegrin and metalloproteinase with thrombospondin motifs 13 | -0.197 | 0.137 | 0.151 | 1 |
| Q7Z7G0 | ABI3BP | Target of Nesh-SH3 | -0.007 | 0.148 | 0.963 | 1 |
| Q7Z7M0 | MEGF8 | Multiple epidermal growth factor-like domains protein 8 | -0.091 | 0.138 | 0.511 | 1 |
| Q86SQ4 | ADGRG6 | Adhesion G-protein coupled receptor G6 | -0.196 | 0.139 | 0.160 | 1 |
| Q86TH1 | ADAMTSL2 | ADAMTS-like protein 2 | -0.186 | 0.128 | 0.145 | 1 |
| Q86U17 | SERPINA11 | Serpin A11 | 0.141 | 0.134 | 0.291 | 1 |
| Q86UD1 | OAF | Out at first protein homolog | 0.078 | 0.137 | 0.572 | 1 |
| Q86UX7 | FERMT3 | Fermitin family homolog 3 | -0.127 | 0.133 | 0.337 | 1 |
| Q86VB7 | CD163 | Scavenger receptor cysteine-rich type 1 protein M130 | -0.078 | 0.135 | 0.564 | 1 |
| Q86YW5 | TREML1 | Trem-like transcript 1 protein | -0.047 | 0.131 | 0.720 | 1 |
| Q86YZ3 | HRNR | Hornerin | 0.028 | 0.116 | 0.811 | 1 |
| Q8IUL8 | CILP2 | Cartilage intermediate layer protein 2 | -0.007 | 0.131 | 0.956 | 1 |
| Q8IZF2 | ADGRF5 | Adhesion G protein-coupled receptor F5 | -0.253 | 0.122 | 0.039 | 1 |
| Q8N1F8 | STK11IP | Serine/threonine-protein kinase 11-interacting protein | -0.067 | 0.119 | 0.573 | 1 |
| Q8N3V7 | SYNPO | Synaptopodin | 0.138 | 0.132 | 0.296 | 1 |
| Q8NBP7 | PCSK9 | Proprotein convertase subtilisin/kexin type 9 | 0.002 | 0.131 | 0.988 | 1 |
| Q8NDA2 | HMCN2 | Hemicentin-2 | -0.069 | 0.124 | 0.577 | 1 |
| Q8WZ75 | ROBO4 | Roundabout homolog 4 | -0.219 | 0.129 | 0.091 | 1 |
| Q92496 | CFHR4 | Complement factor H-related protein 4 | 0.143 | 0.141 | 0.309 | 1 |
| Q92820 | GGH | Gamma-glutamyl hydrolase | -0.129 | 0.127 | 0.310 | 1 |
| Q92954 | PRG4 | Proteoglycan 4 | 0.101 | 0.145 | 0.484 | 1 |
| Q96EE4 | CCDC126 | Coiled-coil domain-containing protein 126 | 0.137 | 0.142 | 0.335 | 1 |
| Q96IY4 | CPB2 | Carboxypeptidase B2 | 0.118 | 0.142 | 0.405 | 1 |
| Q96KG7 | MEGF10 | Multiple epidermal growth factor-like domains protein 10 | 0.137 | 0.113 | 0.223 | 1 |
| Q96KN2 | CNDP1 | Beta-Ala-His dipeptidase | 0.103 | 0.139 | 0.462 | 1 |
| Q96NZ9 | PRAP1 | Proline-rich acidic protein 1 | -0.061 | 0.121 | 0.611 | 1 |
| Q96PD5 | PGLYRP2 | N-acetylmuramoyl-L-alanine amidase | 0.033 | 0.133 | 0.804 | 1 |
| Q96S96 | PEBP4 | Phosphatidylethanolamine-binding protein 4 | 0.243 | 0.146 | 0.095 | 1 |
| Q99784 | OLFM1 | Noelin | -0.134 | 0.136 | 0.323 | 1 |
| Q99969 | RARRES2 | Retinoic acid receptor responder protein 2 | -0.046 | 0.126 | 0.716 | 1 |
| Q9BUN1 | MENT | Protein MENT | 0.090 | 0.113 | 0.424 | 1 |
| Q9BXJ4 | C1QTNF3 | Complement C1q tumor necrosis factor-related protein 3 | 0.016 | 0.129 | 0.899 | 1 |
| Q9BXR6 | CFHR5 | Complement factor H-related protein 5 | 0.153 | 0.140 | 0.277 | 1 |
| Q9BY67 | CADM1 | Cell adhesion molecule 1 | -0.084 | 0.119 | 0.481 | 1 |
| Q9H1U4 | MEGF9 | Multiple epidermal growth factor-like domains protein 9 | 0.022 | 0.142 | 0.875 | 1 |
| Q9H4A9 | DPEP2 | Dipeptidase 2 | -0.020 | 0.137 | 0.884 | 1 |
| Q9H4B7 | TUBB1 | Tubulin beta-1 chain | -0.105 | 0.138 | 0.446 | 1 |
| Q9H4G4 | GLIPR2 | Golgi-associated plant pathogenesis-related protein 1 | -0.285 | 0.128 | 0.025 | 1 |
| Q9H8L6 | MMRN2 | Multimerin-2 | 0.203 | 0.146 | 0.165 | 1 |
| Q9HDC9 | APMAP | Adipocyte plasma membrane-associated protein | -0.065 | 0.133 | 0.628 | 1 |
| Q9NPH3 | IL1RAP | Interleukin-1 receptor accessory protein | 0.097 | 0.135 | 0.472 | 1 |
| Q9NPR2 | SEMA4B | Semaphorin-4B | -0.194 | 0.130 | 0.137 | 1 |
| Q9NPY3 | CD93 | Complement component C1q receptor | 0.060 | 0.134 | 0.653 | 1 |
| Q9NQ79 | CRTAC1 | Cartilage acidic protein 1 | 0.049 | 0.136 | 0.718 | 1 |
| Q9NTU7 | CBLN4 | Cerebellin-4 | 0.082 | 0.153 | 0.589 | 1 |
| Q9NY15 | STAB1 | Stabilin-1 | -0.021 | 0.134 | 0.873 | 1 |
| Q9NY97 | B3GNT2 | N-acetyllactosaminide beta-1,3-N-acetylglucosaminyltransferase 2 | 0.036 | 0.127 | 0.778 | 1 |
| Q9NZK5 | ADA2 | Adenosine deaminase 2 | 0.068 | 0.136 | 0.616 | 1 |
| Q9NZP8 | C1RL | Complement C1r subcomponent-like protein | 0.063 | 0.147 | 0.671 | 1 |
| Q9UBR2 | CTSZ | Cathepsin Z | 0.167 | 0.132 | 0.206 | 1 |
| Q9UBX1 | CTSF | Cathepsin F | -0.094 | 0.124 | 0.447 | 1 |
| Q9UGM5 | FETUB | Fetuin-B | 0.209 | 0.161 | 0.192 | 1 |
| Q9UHG3 | PCYOX1 | Prenylcysteine oxidase 1 | -0.022 | 0.128 | 0.860 | 1 |
| Q9UJJ9 | GNPTG | N-acetylglucosamine-1-phosphotransferase subunit gamma | -0.045 | 0.125 | 0.722 | 1 |
| Q9UK55 | SERPINA10 | Protein Z-dependent protease inhibitor | 0.064 | 0.150 | 0.669 | 1 |
| Q9ULI3 | HEG1 | Protein HEG homolog 1 | 0.011 | 0.125 | 0.932 | 1 |
| Q9UNN8 | PROCR | Endothelial protein C receptor | -0.150 | 0.139 | 0.278 | 1 |
| Q9UNW1 | MINPP1 | Multiple inositol polyphosphate phosphatase 1 | 0.053 | 0.127 | 0.678 | 1 |
| Q9Y251 | HPSE | Heparanase | 0.033 | 0.117 | 0.776 | 1 |
| Q9Y490 | TLN1 | Talin-1 | -0.185 | 0.133 | 0.164 | 1 |
| Q9Y5C1 | ANGPTL3 | Angiopoietin-related protein 3 | 0.026 | 0.135 | 0.846 | 1 |
| Q9Y5Y7 | LYVE1 | Lymphatic vessel endothelial hyaluronic acid receptor 1 | 0.156 | 0.132 | 0.239 | 1 |
| Q9Y646 | CPQ | Carboxypeptidase Q | 0.179 | 0.130 | 0.167 | 1 |
| Q9Y6R7 | FCGBP | IgGFc-binding protein | -0.124 | 0.134 | 0.354 | 1 |
| Q9Y6Z7 | COLEC10 | Collectin-10 | -0.053 | 0.142 | 0.706 | 1 |

**Appendix 4. All 411 plasma proteins of the NECK group in the sensitivity GEE analysis, fully adjusted model.**

| **Protein accession (ID)** | **Gene** | **Protein** | **Estimate** | **Standard error** | **Nominal p-value** | **Holm adjusted p-value** |
| --- | --- | --- | --- | --- | --- | --- |
| A1L4H1 | SSC5D | Soluble scavenger receptor cysteine-rich domain-containing protein SSC5D | 0.071 | 0.142 | 0.616 | 1 |
| O00187 | MASP2 | Mannan-binding lectin serine protease 2 | -0.047 | 0.123 | 0.705 | 1 |
| O00391 | QSOX1 | Sulfhydryl oxidase 1 | -0.169 | 0.141 | 0.231 | 1 |
| O00533 | CHL1 | Neural cell adhesion molecule L1-like protein | -0.149 | 0.130 | 0.253 | 1 |
| O00592 | PODXL | Podocalyxin | 0.217 | 0.145 | 0.136 | 1 |
| O00602 | FCN1 | Ficolin-1 | -0.049 | 0.118 | 0.677 | 1 |
| O14786 | NRP1 | Neuropilin-1 | -0.051 | 0.138 | 0.714 | 1 |
| O14791 | APOL1 | Apolipoprotein L1 | 0.247 | 0.140 | 0.077 | 1 |
| O15204 | ADAMDEC1 | ADAM DEC1 | -0.048 | 0.129 | 0.708 | 1 |
| O43493 | TGOLN2 | Trans-Golgi network integral membrane protein 2 | 0.017 | 0.122 | 0.892 | 1 |
| O43505 | B4GAT1 | Beta-1,4-glucuronyltransferase 1 | 0.269 | 0.179 | 0.132 | 1 |
| O43852 | CALU | Calumenin | -0.133 | 0.117 | 0.257 | 1 |
| O43866 | CD5L | CD5 antigen-like | -0.073 | 0.133 | 0.582 | 1 |
| O75144 | ICOSLG | ICOS ligand | -0.126 | 0.125 | 0.313 | 1 |
| O75594 | PGLYRP1 | Peptidoglycan recognition protein 1 | -0.183 | 0.129 | 0.158 | 1 |
| O75636 | FCN3 | Ficolin-3 | 0.078 | 0.134 | 0.560 | 1 |
| O75882 | ATRN | Attractin | -0.011 | 0.124 | 0.930 | 1 |
| O94985 | CLSTN1 | Calsyntenin-1 | -0.114 | 0.135 | 0.398 | 1 |
| O95445 | APOM | Apolipoprotein M | 0.139 | 0.126 | 0.270 | 1 |
| O95479 | H6PD | GDH/6PGL endoplasmic bifunctional protein | -0.021 | 0.120 | 0.859 | 1 |
| O95497 | VNN1 | Pantetheinase | 0.178 | 0.160 | 0.265 | 1 |
| P00338 | LDHA | L-lactate dehydrogenase A chain | -0.193 | 0.127 | 0.128 | 1 |
| P00441 | SOD1 | Superoxide dismutase [Cu-Zn] | -0.066 | 0.113 | 0.562 | 1 |
| P00450 | CP | Ceruloplasmin | 0.213 | 0.157 | 0.175 | 1 |
| P00488 | F13A1 | Coagulation factor XIII A chain | 0.092 | 0.128 | 0.472 | 1 |
| P00533 | EGFR | Epidermal growth factor receptor | 0.063 | 0.141 | 0.656 | 1 |
| P00734 | F2 | Prothrombin | -0.005 | 0.116 | 0.968 | 1 |
| P00736 | C1R | Complement C1r subcomponent | 0.176 | 0.121 | 0.147 | 1 |
| P00740 | F9 | Coagulation factor IX | 0.355 | 0.156 | 0.023 | 1 |
| P00742 | F10 | Coagulation factor X | 0.113 | 0.132 | 0.391 | 1 |
| P00746 | CFD | Complement factor D | -0.043 | 0.134 | 0.752 | 1 |
| P00747 | PLG | Plasminogen | 0.006 | 0.137 | 0.966 | 1 |
| P00748 | F12 | Coagulation factor XII | 0.066 | 0.133 | 0.618 | 1 |
| P00751 | CFB | Complement factor B | 0.212 | 0.130 | 0.104 | 1 |
| P00915 | CA1 | Carbonic anhydrase 1 | 0.145 | 0.122 | 0.235 | 1 |
| P00918 | CA2 | Carbonic anhydrase 2 | -0.164 | 0.121 | 0.176 | 1 |
| P01008 | SERPINC1 | Antithrombin-III | -0.101 | 0.139 | 0.467 | 1 |
| P01011 | SERPINA3 | Alpha-1-antichymotrypsin | -0.067 | 0.133 | 0.616 | 1 |
| P01019 | AGT | Angiotensinogen | 0.153 | 0.150 | 0.310 | 1 |
| P01024 | C3 | Complement C3 | -0.106 | 0.131 | 0.418 | 1 |
| P01031 | C5 | Complement C5 | 0.254 | 0.124 | 0.040 | 1 |
| P01033 | TIMP1 | Metalloproteinase inhibitor 1 | -0.086 | 0.129 | 0.504 | 1 |
| P01034 | CST3 | Cystatin-C | 0.072 | 0.140 | 0.608 | 1 |
| P01042 | KNG1 | Kininogen-1 | 0.109 | 0.140 | 0.435 | 1 |
| P01137 | TGFB1 | Transforming growth factor beta-1 proprotein | 0.161 | 0.125 | 0.201 | 1 |
| P01344 | IGF2 | Insulin-like growth factor II | 0.240 | 0.134 | 0.073 | 1 |
| P02042 | HBD | Hemoglobin subunit delta | -0.029 | 0.130 | 0.824 | 1 |
| P02144 | MB | Myoglobin | -0.118 | 0.120 | 0.326 | 1 |
| P02452 | COL1A1 | Collagen alpha-1(I) chain | 0.111 | 0.132 | 0.400 | 1 |
| P02533 | KRT14 | Keratin, type I cytoskeletal 14 | -0.038 | 0.118 | 0.748 | 1 |
| P02649 | APOE | Apolipoprotein E | -0.013 | 0.136 | 0.926 | 1 |
| P02652 | APOA2 | Apolipoprotein A-II | 0.045 | 0.130 | 0.730 | 1 |
| P02654 | APOC1 | Apolipoprotein C-I | 0.101 | 0.126 | 0.425 | 1 |
| P02655 | APOC2 | Apolipoprotein C-II | 0.092 | 0.127 | 0.466 | 1 |
| P02656 | APOC3 | Apolipoprotein C-III | 0.115 | 0.134 | 0.389 | 1 |
| P02741 | CRP | C-reactive protein | 0.111 | 0.150 | 0.461 | 1 |
| P02743 | APCS | Serum amyloid P-component | 0.156 | 0.139 | 0.263 | 1 |
| P02745 | C1QA | Complement C1q subcomponent subunit A | 0.141 | 0.127 | 0.266 | 1 |
| P02746 | C1QB | Complement C1q subcomponent subunit B | 0.135 | 0.133 | 0.313 | 1 |
| P02747 | C1QC | Complement C1q subcomponent subunit C | 0.202 | 0.135 | 0.136 | 1 |
| P02748 | C9 | Complement component C9 | -0.166 | 0.135 | 0.221 | 1 |
| P02749 | APOH | Beta-2-glycoprotein 1 | -0.066 | 0.141 | 0.637 | 1 |
| P02750 | LRG1 | Leucine-rich alpha-2-glycoprotein | -0.100 | 0.145 | 0.491 | 1 |
| P02751 | FN1 | Fibronectin | -0.021 | 0.140 | 0.883 | 1 |
| P02753 | RBP4 | Retinol-binding protein 4 | 0.072 | 0.132 | 0.584 | 1 |
| P02760 | AMBP | Protein AMBP | 0.000 | 0.127 | 0.998 | 1 |
| P02765 | AHSG | Alpha-2-HS-glycoprotein | 0.248 | 0.134 | 0.063 | 1 |
| P02766 | TTR | Transthyretin | -0.099 | 0.142 | 0.483 | 1 |
| P02774 | GC | Vitamin D-binding protein | 0.191 | 0.134 | 0.153 | 1 |
| P02775 | PPBP | Platelet basic protein | -0.143 | 0.131 | 0.274 | 1 |
| P02776 | PF4 | Platelet factor 4 | -0.045 | 0.136 | 0.742 | 1 |
| P02790 | HPX | Hemopexin | 0.051 | 0.120 | 0.669 | 1 |
| P03950 | ANG | Angiogenin | -0.031 | 0.131 | 0.813 | 1 |
| P03951 | F11 | Coagulation factor XI | 0.102 | 0.124 | 0.411 | 1 |
| P03952 | KLKB1 | Plasma kallikrein | 0.054 | 0.129 | 0.678 | 1 |
| P03973 | SLPI | Antileukoproteinase | 0.041 | 0.130 | 0.750 | 1 |
| P04003 | C4BPA | C4b-binding protein alpha chain | -0.106 | 0.135 | 0.433 | 1 |
| P04004 | VTN | Vitronectin | 0.331 | 0.136 | 0.015 | 1 |
| P04040 | CAT | Catalase | 0.215 | 0.125 | 0.085 | 1 |
| P04070 | PROC | Vitamin K-dependent protein C | 0.046 | 0.127 | 0.718 | 1 |
| P04075 | ALDOA | Fructose-bisphosphate aldolase A | 0.013 | 0.141 | 0.928 | 1 |
| P04114 | APOB | Apolipoprotein B-100 | -0.273 | 0.133 | 0.039 | 1 |
| P04180 | LCAT | Phosphatidylcholine-sterol acyltransferase | 0.040 | 0.133 | 0.763 | 1 |
| P04196 | HRG | Histidine-rich glycoprotein | 0.048 | 0.140 | 0.734 | 1 |
| P04217 | A1BG | Alpha-1B-glycoprotein | 0.049 | 0.136 | 0.720 | 1 |
| P04259 | KRT6B | Keratin, type II cytoskeletal 6B | 0.091 | 0.132 | 0.494 | 1 |
| P04264 | KRT1 | Keratin, type II cytoskeletal 1 | -0.043 | 0.114 | 0.704 | 1 |
| P04275 | VWF | von Willebrand factor | -0.032 | 0.130 | 0.805 | 1 |
| P04278 | SHBG | Sex hormone-binding globulin | 0.146 | 0.179 | 0.415 | 1 |
| P04406 | GAPDH | Glyceraldehyde-3-phosphate dehydrogenase | 0.014 | 0.120 | 0.906 | 1 |
| P04439 | HLA-A | HLA class I histocompatibility antigen, A alpha chain | -0.069 | 0.131 | 0.602 | 1 |
| P04746 | AMY2A | Pancreatic alpha-amylase | 0.220 | 0.131 | 0.092 | 1 |
| P05019 | IGF1 | Insulin-like growth factor I | -0.046 | 0.129 | 0.721 | 1 |
| P05062 | ALDOB | Fructose-bisphosphate aldolase B | -0.119 | 0.146 | 0.415 | 1 |
| P05067 | APP | Amyloid-beta precursor protein | 0.015 | 0.118 | 0.900 | 1 |
| P05090 | APOD | Apolipoprotein D | -0.039 | 0.127 | 0.759 | 1 |
| P05106 | ITGB3 | Integrin beta-3 | -0.001 | 0.123 | 0.994 | 1 |
| P05109 | S100A8 | Protein S100-A8 | 0.065 | 0.126 | 0.606 | 1 |
| P05121 | SERPINE1 | Plasminogen activator inhibitor 1 | -0.170 | 0.119 | 0.152 | 1 |
| P05154 | SERPINA5 | Plasma serine protease inhibitor | 0.049 | 0.130 | 0.708 | 1 |
| P05155 | SERPING1 | Plasma protease C1 inhibitor | 0.036 | 0.126 | 0.775 | 1 |
| P05156 | CFI | Complement factor I | 0.245 | 0.132 | 0.063 | 1 |
| P05160 | F13B | Coagulation factor XIII B chain | 0.074 | 0.128 | 0.563 | 1 |
| P05164 | MPO | Myeloperoxidase | -0.049 | 0.122 | 0.686 | 1 |
| P05362 | ICAM1 | Intercellular adhesion molecule 1 | -0.086 | 0.125 | 0.490 | 1 |
| P05451 | REG1A | Lithostathine-1-alpha | -0.111 | 0.118 | 0.349 | 1 |
| P05452 | CLEC3B | Tetranectin | 0.022 | 0.142 | 0.876 | 1 |
| P05543 | SERPINA7 | Thyroxine-binding globulin | 0.348 | 0.154 | 0.024 | 1 |
| P05546 | SERPIND1 | Heparin cofactor 2 | 0.102 | 0.142 | 0.474 | 1 |
| P05556 | ITGB1 | Integrin beta-1 | -0.266 | 0.123 | 0.031 | 1 |
| P06276 | BCHE | Cholinesterase | -0.129 | 0.163 | 0.430 | 1 |
| P06396 | GSN | Gelsolin | 0.110 | 0.133 | 0.410 | 1 |
| P06681 | C2 | Complement C2 | -0.017 | 0.127 | 0.891 | 1 |
| P06702 | S100A9 | Protein S100-A9 | -0.096 | 0.134 | 0.474 | 1 |
| P06727 | APOA4 | Apolipoprotein A-IV | 0.027 | 0.126 | 0.834 | 1 |
| P06732 | CKM | Creatine kinase M-type | 0.184 | 0.146 | 0.210 | 1 |
| P07195 | LDHB | L-lactate dehydrogenase B chain | 0.133 | 0.126 | 0.290 | 1 |
| P07225 | PROS1 | Vitamin K-dependent protein S | 0.084 | 0.142 | 0.555 | 1 |
| P07237 | P4HB | Protein disulfide-isomerase | -0.085 | 0.134 | 0.526 | 1 |
| P07333 | CSF1R | Macrophage colony-stimulating factor 1 receptor | -0.162 | 0.127 | 0.201 | 1 |
| P07339 | CTSD | Cathepsin D | -0.042 | 0.116 | 0.718 | 1 |
| P07357 | C8A | Complement component C8 alpha chain | 0.167 | 0.135 | 0.215 | 1 |
| P07358 | C8B | Complement component C8 beta chain | 0.175 | 0.123 | 0.156 | 1 |
| P07359 | GP1BA | Platelet glycoprotein Ib alpha chain | -0.021 | 0.128 | 0.871 | 1 |
| P07360 | C8G | Complement component C8 gamma chain | 0.245 | 0.131 | 0.062 | 1 |
| P07437 | TUBB | Tubulin beta chain | 0.093 | 0.135 | 0.492 | 1 |
| P07602 | PSAP | Prosaposin | -0.094 | 0.131 | 0.471 | 1 |
| P07737 | PFN1 | Profilin-1 | -0.180 | 0.138 | 0.194 | 1 |
| P07911 | UMOD | Uromodulin | 0.114 | 0.133 | 0.390 | 1 |
| P07942 | LAMB1 | Laminin subunit beta-1 | 0.041 | 0.124 | 0.742 | 1 |
| P07951 | TPM2 | Tropomyosin beta chain | -0.122 | 0.121 | 0.310 | 1 |
| P07996 | THBS1 | Thrombospondin-1 | -0.007 | 0.126 | 0.959 | 1 |
| P07998 | RNASE1 | Ribonuclease pancreatic | -0.023 | 0.125 | 0.857 | 1 |
| P08185 | SERPINA6 | Corticosteroid-binding globulin | 0.154 | 0.152 | 0.312 | 1 |
| P08195 | SLC3A2 | 4F2 cell-surface antigen heavy chain | 0.069 | 0.139 | 0.619 | 1 |
| P08238 | HSP90AB1 | Heat shock protein HSP 90-beta | -0.025 | 0.115 | 0.829 | 1 |
| P08253 | MMP2 | 72 kDa type IV collagenase | -0.034 | 0.130 | 0.792 | 1 |
| P08294 | SOD3 | Extracellular superoxide dismutase [Cu-Zn] | 0.021 | 0.112 | 0.849 | 1 |
| P08311 | CTSG | Cathepsin G | -0.166 | 0.119 | 0.165 | 1 |
| P08493 | MGP | Matrix Gla protein | 0.184 | 0.145 | 0.205 | 1 |
| P08514 | ITGA2B | Integrin alpha-IIb | -0.063 | 0.130 | 0.626 | 1 |
| P08519 | LPA | Apolipoprotein(a) | -0.090 | 0.123 | 0.462 | 1 |
| P08571 | CD14 | Monocyte differentiation antigen CD14 | 0.227 | 0.132 | 0.086 | 1 |
| P08603 | CFH | Complement factor H | -0.017 | 0.134 | 0.902 | 1 |
| P08697 | SERPINF2 | Alpha-2-antiplasmin | 0.263 | 0.129 | 0.042 | 1 |
| P08709 | F7 | Coagulation factor VII | 0.153 | 0.139 | 0.269 | 1 |
| P08779 | KRT16 | Keratin, type I cytoskeletal 16 | 0.153 | 0.120 | 0.203 | 1 |
| P08887 | IL6R | Interleukin-6 receptor subunit alpha | 0.033 | 0.123 | 0.787 | 1 |
| P09172 | DBH | Dopamine beta-hydroxylase | 0.229 | 0.134 | 0.086 | 1 |
| P09486 | SPARC | SPARC | -0.313 | 0.144 | 0.029 | 1 |
| P09871 | C1S | Complement C1s subcomponent | 0.017 | 0.123 | 0.890 | 1 |
| P09960 | LTA4H | Leukotriene A-4 hydrolase | 0.205 | 0.127 | 0.106 | 1 |
| P0C0L4 | C4A | Complement C4-A | 0.012 | 0.131 | 0.926 | 1 |
| P0C0L5 | C4B | Complement C4-B | 0.423 | 0.135 | 0.002 | 0.703257 |
| P0DJI8 | SAA1 | Serum amyloid A-1 protein | 0.010 | 0.133 | 0.940 | 1 |
| P10124 | SRGN | Serglycin | -0.020 | 0.119 | 0.865 | 1 |
| P10153 | RNASE2 | Non-secretory ribonuclease | -0.236 | 0.125 | 0.058 | 1 |
| P10586 | PTPRF | Receptor-type tyrosine-protein phosphatase F | 0.034 | 0.116 | 0.768 | 1 |
| P10643 | C7 | Complement component C7 | 0.212 | 0.135 | 0.117 | 1 |
| P10646 | TFPI | Tissue factor pathway inhibitor | -0.179 | 0.125 | 0.153 | 1 |
| P10720 | PF4V1 | Platelet factor 4 variant | 0.081 | 0.127 | 0.523 | 1 |
| P10721 | KIT | Mast/stem cell growth factor receptor Kit | -0.106 | 0.125 | 0.396 | 1 |
| P10909 | CLU | Clusterin | -0.014 | 0.124 | 0.911 | 1 |
| P11021 | HSPA5 | Endoplasmic reticulum chaperone BiP | -0.131 | 0.132 | 0.321 | 1 |
| P11142 | HSPA8 | Heat shock cognate 71 kDa protein | -0.110 | 0.122 | 0.368 | 1 |
| P11226 | MBL2 | Mannose-binding protein C | -0.186 | 0.123 | 0.130 | 1 |
| P11279 | LAMP1 | Lysosome-associated membrane glycoprotein 1 | 0.071 | 0.137 | 0.601 | 1 |
| P11362 | FGFR1 | Fibroblast growth factor receptor 1 | -0.040 | 0.123 | 0.746 | 1 |
| P11597 | CETP | Cholesteryl ester transfer protein | 0.092 | 0.128 | 0.476 | 1 |
| P11717 | IGF2R | Cation-independent mannose-6-phosphate receptor | -0.172 | 0.120 | 0.152 | 1 |
| P12109 | COL6A1 | Collagen alpha-1(VI) chain | -0.150 | 0.124 | 0.227 | 1 |
| P12111 | COL6A3 | Collagen alpha-3(VI) chain | 0.016 | 0.133 | 0.902 | 1 |
| P12259 | F5 | Coagulation factor V | 0.095 | 0.122 | 0.435 | 1 |
| P12821 | ACE | Angiotensin-converting enzyme | -0.126 | 0.120 | 0.292 | 1 |
| P12830 | CDH1 | Cadherin-1 | -0.215 | 0.121 | 0.075 | 1 |
| P12955 | PEPD | Xaa-Pro dipeptidase | -0.130 | 0.134 | 0.330 | 1 |
| P13473 | LAMP2 | Lysosome-associated membrane glycoprotein 2 | 0.074 | 0.134 | 0.583 | 1 |
| P13591 | NCAM1 | Neural cell adhesion molecule 1 | -0.128 | 0.164 | 0.436 | 1 |
| P13598 | ICAM2 | Intercellular adhesion molecule 2 | 0.090 | 0.140 | 0.520 | 1 |
| P13645 | KRT10 | Keratin, type I cytoskeletal 10 | 0.014 | 0.118 | 0.905 | 1 |
| P13646 | KRT13 | Keratin, type I cytoskeletal 13 | -0.077 | 0.128 | 0.548 | 1 |
| P13671 | C6 | Complement component C6 | 0.112 | 0.131 | 0.392 | 1 |
| P13727 | PRG2 | Bone marrow proteoglycan | 0.033 | 0.132 | 0.802 | 1 |
| P13796 | LCP1 | Plastin-2 | 0.103 | 0.127 | 0.419 | 1 |
| P14151 | SELL | L-selectin | 0.021 | 0.129 | 0.870 | 1 |
| P14209 | CD99 | CD99 antigen | 0.222 | 0.142 | 0.120 | 1 |
| P14314 | PRKCSH | Glucosidase 2 subunit beta | 0.095 | 0.109 | 0.380 | 1 |
| P14543 | NID1 | Nidogen-1 | 0.019 | 0.127 | 0.879 | 1 |
| P14618 | PKM | Pyruvate kinase PKM | 0.066 | 0.129 | 0.609 | 1 |
| P14625 | HSP90B1 | Endoplasmin | 0.032 | 0.130 | 0.803 | 1 |
| P14780 | MMP9 | Matrix metalloproteinase-9 | 0.056 | 0.126 | 0.658 | 1 |
| P15144 | ANPEP | Aminopeptidase N | -0.185 | 0.130 | 0.155 | 1 |
| P15151 | PVR | Poliovirus receptor | 0.002 | 0.129 | 0.985 | 1 |
| P15169 | CPN1 | Carboxypeptidase N catalytic chain | -0.023 | 0.145 | 0.875 | 1 |
| P16035 | TIMP2 | Metalloproteinase inhibitor 2 | 0.226 | 0.125 | 0.072 | 1 |
| P16070 | CD44 | CD44 antigen | 0.020 | 0.123 | 0.873 | 1 |
| P16112 | ACAN | Aggrecan core protein | 0.061 | 0.122 | 0.621 | 1 |
| P16930 | FAH | Fumarylacetoacetase | -0.021 | 0.129 | 0.868 | 1 |
| P17813 | ENG | Endoglin | -0.038 | 0.120 | 0.753 | 1 |
| P17936 | IGFBP3 | Insulin-like growth factor-binding protein 3 | 0.148 | 0.132 | 0.260 | 1 |
| P18065 | IGFBP2 | Insulin-like growth factor-binding protein 2 | 0.053 | 0.137 | 0.701 | 1 |
| P18206 | VCL | Vinculin | -0.233 | 0.131 | 0.076 | 1 |
| P18428 | LBP | Lipopolysaccharide-binding protein | 0.049 | 0.131 | 0.709 | 1 |
| P19021 | PAM | Peptidyl-glycine alpha-amidating monooxygenase | 0.046 | 0.138 | 0.742 | 1 |
| P19022 | CDH2 | Cadherin-2 | 0.163 | 0.126 | 0.194 | 1 |
| P19320 | VCAM1 | Vascular cell adhesion protein 1 | -0.055 | 0.133 | 0.676 | 1 |
| P19823 | ITIH2 | Inter-alpha-trypsin inhibitor heavy chain H2 | 0.071 | 0.126 | 0.574 | 1 |
| P19827 | ITIH1 | Inter-alpha-trypsin inhibitor heavy chain H1 | 0.250 | 0.144 | 0.083 | 1 |
| P20023 | CR2 | Complement receptor type 2 | 0.098 | 0.148 | 0.507 | 1 |
| P20742 | PZP | Pregnancy zone protein | 0.117 | 0.161 | 0.466 | 1 |
| P20851 | C4BPB | C4b-binding protein beta chain | 0.050 | 0.134 | 0.709 | 1 |
| P21333 | FLNA | Filamin-A | -0.040 | 0.113 | 0.720 | 1 |
| P22105 | TNXB | Tenascin-X | -0.175 | 0.126 | 0.167 | 1 |
| P22352 | GPX3 | Glutathione peroxidase 3 | -0.131 | 0.135 | 0.333 | 1 |
| P22692 | IGFBP4 | Insulin-like growth factor-binding protein 4 | -0.052 | 0.135 | 0.700 | 1 |
| P22792 | CPN2 | Carboxypeptidase N subunit 2 | 0.060 | 0.142 | 0.675 | 1 |
| P22891 | PROZ | Vitamin K-dependent protein Z | 0.125 | 0.129 | 0.332 | 1 |
| P22897 | MRC1 | Macrophage mannose receptor 1 | -0.031 | 0.119 | 0.797 | 1 |
| P23142 | FBLN1 | Fibulin-1 | 0.251 | 0.119 | 0.035 | 1 |
| P23470 | PTPRG | Receptor-type tyrosine-protein phosphatase gamma | 0.076 | 0.127 | 0.550 | 1 |
| P23528 | CFL1 | Cofilin-1 | -0.063 | 0.125 | 0.612 | 1 |
| P24043 | LAMA2 | Laminin subunit alpha-2 | 0.256 | 0.142 | 0.072 | 1 |
| P24592 | IGFBP6 | Insulin-like growth factor-binding protein 6 | 0.254 | 0.128 | 0.047 | 1 |
| P24593 | IGFBP5 | Insulin-like growth factor-binding protein 5 | 0.244 | 0.142 | 0.085 | 1 |
| P24821 | TNC | Tenascin | -0.064 | 0.121 | 0.599 | 1 |
| P25311 | AZGP1 | Zinc-alpha-2-glycoprotein | 0.180 | 0.151 | 0.235 | 1 |
| P25774 | CTSS | Cathepsin S | 0.017 | 0.130 | 0.894 | 1 |
| P26038 | MSN | Moesin | -0.076 | 0.133 | 0.568 | 1 |
| P26927 | MST1 | Hepatocyte growth factor-like protein | -0.166 | 0.107 | 0.122 | 1 |
| P27169 | PON1 | Serum paraoxonase/arylesterase 1 | 0.141 | 0.134 | 0.291 | 1 |
| P27487 | DPP4 | Dipeptidyl peptidase 4 | -0.228 | 0.135 | 0.091 | 1 |
| P27797 | CALR | Calreticulin | -0.200 | 0.126 | 0.112 | 1 |
| P27918 | CFP | Properdin | 0.128 | 0.132 | 0.331 | 1 |
| P28799 | GRN | Progranulin | 0.082 | 0.140 | 0.558 | 1 |
| P29401 | TKT | Transketolase | 0.065 | 0.136 | 0.633 | 1 |
| P29622 | SERPINA4 | Kallistatin | 0.033 | 0.132 | 0.804 | 1 |
| P30043 | BLVRB | Flavin reductase (NADPH) | -0.031 | 0.120 | 0.794 | 1 |
| P30101 | PDIA3 | Protein disulfide-isomerase A3 | 0.197 | 0.141 | 0.162 | 1 |
| P32119 | PRDX2 | Peroxiredoxin-2 | 0.050 | 0.124 | 0.688 | 1 |
| P32942 | ICAM3 | Intercellular adhesion molecule 3 | -0.137 | 0.124 | 0.267 | 1 |
| P33151 | CDH5 | Cadherin-5 | 0.097 | 0.140 | 0.488 | 1 |
| P33908 | MAN1A1 | Mannosyl-oligosaccharide 1,2-alpha-mannosidase IA | -0.012 | 0.131 | 0.929 | 1 |
| P34096 | RNASE4 | Ribonuclease 4 | -0.013 | 0.134 | 0.923 | 1 |
| P35030 | PRSS3 | Trypsin-3 | -0.318 | 0.133 | 0.017 | 1 |
| P35443 | THBS4 | Thrombospondin-4 | 0.007 | 0.135 | 0.958 | 1 |
| P35527 | KRT9 | Keratin, type I cytoskeletal 9 | 0.098 | 0.113 | 0.382 | 1 |
| P35542 | SAA4 | Serum amyloid A-4 protein | -0.079 | 0.132 | 0.548 | 1 |
| P35555 | FBN1 | Fibrillin-1 | 0.039 | 0.126 | 0.754 | 1 |
| P35858 | IGFALS | Insulin-like growth factor-binding protein complex acid labile subunit | 0.184 | 0.134 | 0.171 | 1 |
| P35908 | KRT2 | Keratin, type II cytoskeletal 2 epidermal | -0.078 | 0.114 | 0.496 | 1 |
| P35916 | FLT4 | Vascular endothelial growth factor receptor 3 | 0.188 | 0.136 | 0.166 | 1 |
| P36955 | SERPINF1 | Pigment epithelium-derived factor | 0.372 | 0.137 | 0.007 | 1 |
| P36980 | CFHR2 | Complement factor H-related protein 2 | 0.185 | 0.142 | 0.194 | 1 |
| P39060 | COL18A1 | Collagen alpha-1(XVIII) chain | 0.195 | 0.142 | 0.169 | 1 |
| P40189 | IL6ST | Interleukin-6 receptor subunit beta | -0.158 | 0.135 | 0.239 | 1 |
| P40197 | GP5 | Platelet glycoprotein V | -0.149 | 0.128 | 0.246 | 1 |
| P41222 | PTGDS | Prostaglandin-H2 D-isomerase | 0.087 | 0.129 | 0.502 | 1 |
| P43121 | MCAM | Cell surface glycoprotein MUC18 | 0.182 | 0.126 | 0.149 | 1 |
| P43251 | BTD | Biotinidase | 0.036 | 0.133 | 0.786 | 1 |
| P43652 | AFM | Afamin | -0.012 | 0.119 | 0.918 | 1 |
| P48594 | SERPINB4 | Serpin B4 | -0.133 | 0.120 | 0.266 | 1 |
| P48668 | KRT6C | Keratin, type II cytoskeletal 6C | -0.055 | 0.115 | 0.636 | 1 |
| P48740 | MASP1 | Mannan-binding lectin serine protease 1 | -0.115 | 0.127 | 0.366 | 1 |
| P49747 | COMP | Cartilage oligomeric matrix protein | 0.089 | 0.126 | 0.482 | 1 |
| P49908 | SELENOP | Selenoprotein P | -0.003 | 0.141 | 0.983 | 1 |
| P49913 | CAMP | Cathelicidin antimicrobial peptide | 0.071 | 0.134 | 0.593 | 1 |
| P50395 | GDI2 | Rab GDP dissociation inhibitor beta | -0.061 | 0.129 | 0.637 | 1 |
| P51884 | LUM | Lumican | 0.133 | 0.137 | 0.331 | 1 |
| P53634 | CTSC | Dipeptidyl peptidase 1 | -0.057 | 0.118 | 0.631 | 1 |
| P54289 | CACNA2D1 | Voltage-dependent calcium channel subunit alpha-2/delta-1 | 0.195 | 0.126 | 0.122 | 1 |
| P54802 | NAGLU | Alpha-N-acetylglucosaminidase | 0.062 | 0.132 | 0.639 | 1 |
| P55056 | APOC4 | Apolipoprotein C-IV | 0.070 | 0.141 | 0.619 | 1 |
| P55058 | PLTP | Phospholipid transfer protein | 0.056 | 0.129 | 0.663 | 1 |
| P55103 | INHBC | Inhibin beta C chain | 0.043 | 0.118 | 0.713 | 1 |
| P55290 | CDH13 | Cadherin-13 | -0.041 | 0.130 | 0.751 | 1 |
| P59666 | DEFA3 | Neutrophil defensin 3 | 0.091 | 0.126 | 0.473 | 1 |
| P60174 | TPI1 | Triosephosphate isomerase | 0.083 | 0.117 | 0.475 | 1 |
| P60709 | ACTB | Actin, cytoplasmic 1 | 0.030 | 0.123 | 0.809 | 1 |
| P61224 | RAP1B | Ras-related protein Rap-1b | -0.056 | 0.131 | 0.667 | 1 |
| P61626 | LYZ | Lysozyme C | 0.099 | 0.137 | 0.473 | 1 |
| P61769 | B2M | Beta-2-microglobulin | 0.136 | 0.129 | 0.292 | 1 |
| P62328 | TMSB4X | Thymosin beta-4 | -0.235 | 0.125 | 0.060 | 1 |
| P62937 | PPIA | Peptidyl-prolyl cis-trans isomerase A | -0.208 | 0.127 | 0.101 | 1 |
| P63104 | YWHAZ | 14-3-3 protein zeta/delta | 0.080 | 0.132 | 0.543 | 1 |
| P63261 | ACTG1 | Actin, cytoplasmic 2 | -0.063 | 0.133 | 0.638 | 1 |
| P68032.P68133 | ACTC1;ACTA1 | Actin, alpha cardiac muscle 1;Actin, alpha skeletal muscle | 0.025 | 0.128 | 0.847 | 1 |
| P68363 | TUBA1B | Tubulin alpha-1B chain | 0.093 | 0.124 | 0.453 | 1 |
| P68871 | HBB | Hemoglobin subunit beta | -0.063 | 0.139 | 0.652 | 1 |
| P69892 | HBG2 | Hemoglobin subunit gamma-2 | -0.127 | 0.120 | 0.291 | 1 |
| P69905 | HBA1 | Hemoglobin subunit alpha | 0.002 | 0.129 | 0.987 | 1 |
| P78509 | RELN | Reelin | 0.246 | 0.130 | 0.058 | 1 |
| P80108 | GPLD1 | Phosphatidylinositol-glycan-specific phospholipase D | -0.098 | 0.140 | 0.481 | 1 |
| P80723 | BASP1 | Brain acid soluble protein 1 | 0.068 | 0.123 | 0.577 | 1 |
| P98160 | HSPG2 | Basement membrane-specific heparan sulfate proteoglycan core protein | 0.053 | 0.142 | 0.710 | 1 |
| Q00610 | CLTC | Clathrin heavy chain 1 | 0.019 | 0.124 | 0.877 | 1 |
| Q01459 | CTBS | Di-N-acetylchitobiase | -0.204 | 0.128 | 0.109 | 1 |
| Q02985 | CFHR3 | Complement factor H-related protein 3 | 0.086 | 0.131 | 0.510 | 1 |
| Q03591 | CFHR1 | Complement factor H-related protein 1 | -0.092 | 0.127 | 0.470 | 1 |
| Q04721 | NOTCH2 | Neurogenic locus notch homolog protein 2 | -0.089 | 0.128 | 0.486 | 1 |
| Q04756 | HGFAC | Hepatocyte growth factor activator | 0.049 | 0.129 | 0.701 | 1 |
| Q06033 | ITIH3 | Inter-alpha-trypsin inhibitor heavy chain H3 | -0.128 | 0.130 | 0.322 | 1 |
| Q06830 | PRDX1 | Peroxiredoxin-1 | 0.157 | 0.128 | 0.220 | 1 |
| Q07954 | LRP1 | Prolow-density lipoprotein receptor-related protein 1 | -0.140 | 0.128 | 0.274 | 1 |
| Q08380 | LGALS3BP | Galectin-3-binding protein | 0.082 | 0.147 | 0.577 | 1 |
| Q10588 | BST1 | ADP-ribosyl cyclase/cyclic ADP-ribose hydrolase 2 | 0.189 | 0.150 | 0.209 | 1 |
| Q12805 | EFEMP1 | EGF-containing fibulin-like extracellular matrix protein 1 | 0.218 | 0.136 | 0.107 | 1 |
| Q12841 | FSTL1 | Follistatin-related protein 1 | -0.040 | 0.133 | 0.764 | 1 |
| Q12860 | CNTN1 | Contactin-1 | -0.087 | 0.127 | 0.492 | 1 |
| Q12884 | FAP | Prolyl endopeptidase FAP | -0.038 | 0.124 | 0.759 | 1 |
| Q12907 | LMAN2 | Vesicular integral-membrane protein VIP36 | 0.044 | 0.141 | 0.753 | 1 |
| Q12913 | PTPRJ | Receptor-type tyrosine-protein phosphatase eta | 0.148 | 0.139 | 0.287 | 1 |
| Q13093 | PLA2G7 | Platelet-activating factor acetylhydrolase | -0.036 | 0.135 | 0.789 | 1 |
| Q13103 | SPP2 | Secreted phosphoprotein 24 | 0.069 | 0.152 | 0.648 | 1 |
| Q13201 | MMRN1 | Multimerin-1 | -0.206 | 0.120 | 0.086 | 1 |
| Q13332 | PTPRS | Receptor-type tyrosine-protein phosphatase S | 0.035 | 0.137 | 0.796 | 1 |
| Q13449 | LSAMP | Limbic system-associated membrane protein | 0.043 | 0.123 | 0.725 | 1 |
| Q13740 | ALCAM | CD166 antigen | -0.034 | 0.134 | 0.800 | 1 |
| Q13790 | APOF | Apolipoprotein F | -0.084 | 0.126 | 0.507 | 1 |
| Q13822 | ENPP2 | Ectonucleotide pyrophosphatase/phosphodiesterase family member 2 | 0.156 | 0.137 | 0.254 | 1 |
| Q14112 | NID2 | Nidogen-2 | 0.088 | 0.115 | 0.443 | 1 |
| Q14126 | DSG2 | Desmoglein-2 | 0.177 | 0.146 | 0.227 | 1 |
| Q14314 | FGL2 | Fibroleukin | -0.050 | 0.114 | 0.662 | 1 |
| Q14515 | SPARCL1 | SPARC-like protein 1 | 0.035 | 0.125 | 0.781 | 1 |
| Q14520 | HABP2 | Hyaluronan-binding protein 2 | -0.038 | 0.134 | 0.775 | 1 |
| Q14624 | ITIH4 | Inter-alpha-trypsin inhibitor heavy chain H4 | 0.182 | 0.146 | 0.214 | 1 |
| Q14766 | LTBP1 | Latent-transforming growth factor beta-binding protein 1 | -0.019 | 0.131 | 0.887 | 1 |
| Q14956 | GPNMB | Transmembrane glycoprotein NMB | -0.142 | 0.138 | 0.304 | 1 |
| Q15063 | POSTN | Periostin | -0.058 | 0.123 | 0.636 | 1 |
| Q15113 | PCOLCE | Procollagen C-endopeptidase enhancer 1 | -0.146 | 0.122 | 0.230 | 1 |
| Q15166 | PON3 | Serum paraoxonase/lactonase 3 | 0.011 | 0.125 | 0.929 | 1 |
| Q15485 | FCN2 | Ficolin-2 | -0.153 | 0.140 | 0.272 | 1 |
| Q15582 | TGFBI | Transforming growth factor-beta-induced protein ig-h3 | 0.013 | 0.129 | 0.917 | 1 |
| Q15828 | CST6 | Cystatin-M | -0.040 | 0.123 | 0.744 | 1 |
| Q15848 | ADIPOQ | Adiponectin | 0.182 | 0.133 | 0.172 | 1 |
| Q16270 | IGFBP7 | Insulin-like growth factor-binding protein 7 | -0.043 | 0.126 | 0.733 | 1 |
| Q16610 | ECM1 | Extracellular matrix protein 1 | 0.187 | 0.138 | 0.175 | 1 |
| Q16706 | MAN2A1 | Alpha-mannosidase 2 | 0.058 | 0.133 | 0.665 | 1 |
| Q16853 | AOC3 | Membrane primary amine oxidase | -0.086 | 0.121 | 0.478 | 1 |
| Q4LDE5 | SVEP1 | Sushi, von Willebrand factor type A, EGF and pentraxin domain-containing protein 1 | -0.012 | 0.115 | 0.913 | 1 |
| Q562R1 | ACTBL2 | Beta-actin-like protein 2 | -0.164 | 0.114 | 0.149 | 1 |
| Q6EMK4 | VASN | Vasorin | 0.020 | 0.146 | 0.894 | 1 |
| Q6UWP8 | SBSN | Suprabasin | -0.082 | 0.125 | 0.513 | 1 |
| Q6UX71 | PLXDC2 | Plexin domain-containing protein 2 | 0.072 | 0.130 | 0.579 | 1 |
| Q6UXB8 | PI16 | Peptidase inhibitor 16 | -0.127 | 0.133 | 0.339 | 1 |
| Q6UY14 | ADAMTSL4 | ADAMTS-like protein 4 | 0.029 | 0.132 | 0.826 | 1 |
| Q6YHK3 | CD109 | CD109 antigen | -0.033 | 0.133 | 0.805 | 1 |
| Q76LX8 | ADAMTS13 | A disintegrin and metalloproteinase with thrombospondin motifs 13 | 0.190 | 0.131 | 0.148 | 1 |
| Q7Z7G0 | ABI3BP | Target of Nesh-SH3 | 0.063 | 0.145 | 0.664 | 1 |
| Q7Z7M0 | MEGF8 | Multiple epidermal growth factor-like domains protein 8 | 0.054 | 0.131 | 0.681 | 1 |
| Q86SQ4 | ADGRG6 | Adhesion G-protein coupled receptor G6 | -0.032 | 0.142 | 0.822 | 1 |
| Q86TH1 | ADAMTSL2 | ADAMTS-like protein 2 | 0.010 | 0.109 | 0.928 | 1 |
| Q86U17 | SERPINA11 | Serpin A11 | 0.069 | 0.150 | 0.646 | 1 |
| Q86UD1 | OAF | Out at first protein homolog | 0.013 | 0.134 | 0.925 | 1 |
| Q86UX7 | FERMT3 | Fermitin family homolog 3 | -0.168 | 0.120 | 0.161 | 1 |
| Q86VB7 | CD163 | Scavenger receptor cysteine-rich type 1 protein M130 | -0.048 | 0.127 | 0.707 | 1 |
| Q86YW5 | TREML1 | Trem-like transcript 1 protein | -0.092 | 0.113 | 0.417 | 1 |
| Q86YZ3 | HRNR | Hornerin | 0.125 | 0.123 | 0.310 | 1 |
| Q8IUL8 | CILP2 | Cartilage intermediate layer protein 2 | 0.036 | 0.126 | 0.778 | 1 |
| Q8IZF2 | ADGRF5 | Adhesion G protein-coupled receptor F5 | 0.171 | 0.117 | 0.143 | 1 |
| Q8N1F8 | STK11IP | Serine/threonine-protein kinase 11-interacting protein | 0.143 | 0.149 | 0.336 | 1 |
| Q8N3V7 | SYNPO | Synaptopodin | 0.136 | 0.126 | 0.283 | 1 |
| Q8NBP7 | PCSK9 | Proprotein convertase subtilisin/kexin type 9 | -0.072 | 0.132 | 0.585 | 1 |
| Q8NDA2 | HMCN2 | Hemicentin-2 | -0.123 | 0.117 | 0.296 | 1 |
| Q8WZ75 | ROBO4 | Roundabout homolog 4 | 0.134 | 0.139 | 0.336 | 1 |
| Q92496 | CFHR4 | Complement factor H-related protein 4 | 0.008 | 0.126 | 0.951 | 1 |
| Q92820 | GGH | Gamma-glutamyl hydrolase | -0.049 | 0.135 | 0.719 | 1 |
| Q92954 | PRG4 | Proteoglycan 4 | 0.017 | 0.138 | 0.903 | 1 |
| Q96EE4 | CCDC126 | Coiled-coil domain-containing protein 126 | 0.025 | 0.121 | 0.836 | 1 |
| Q96IY4 | CPB2 | Carboxypeptidase B2 | 0.191 | 0.129 | 0.140 | 1 |
| Q96KG7 | MEGF10 | Multiple epidermal growth factor-like domains protein 10 | -0.238 | 0.112 | 0.033 | 1 |
| Q96KN2 | CNDP1 | Beta-Ala-His dipeptidase | 0.075 | 0.132 | 0.570 | 1 |
| Q96NZ9 | PRAP1 | Proline-rich acidic protein 1 | -0.219 | 0.118 | 0.063 | 1 |
| Q96PD5 | PGLYRP2 | N-acetylmuramoyl-L-alanine amidase | 0.000 | 0.131 | 0.999 | 1 |
| Q96S96 | PEBP4 | Phosphatidylethanolamine-binding protein 4 | 0.051 | 0.150 | 0.737 | 1 |
| Q99784 | OLFM1 | Noelin | -0.238 | 0.123 | 0.054 | 1 |
| Q99969 | RARRES2 | Retinoic acid receptor responder protein 2 | -0.135 | 0.118 | 0.252 | 1 |
| Q9BUN1 | MENT | Protein MENT | 0.051 | 0.108 | 0.637 | 1 |
| Q9BXJ4 | C1QTNF3 | Complement C1q tumor necrosis factor-related protein 3 | -0.220 | 0.119 | 0.066 | 1 |
| Q9BXR6 | CFHR5 | Complement factor H-related protein 5 | 0.161 | 0.130 | 0.214 | 1 |
| Q9BY67 | CADM1 | Cell adhesion molecule 1 | -0.089 | 0.116 | 0.447 | 1 |
| Q9H1U4 | MEGF9 | Multiple epidermal growth factor-like domains protein 9 | -0.067 | 0.133 | 0.616 | 1 |
| Q9H4A9 | DPEP2 | Dipeptidase 2 | 0.099 | 0.124 | 0.425 | 1 |
| Q9H4B7 | TUBB1 | Tubulin beta-1 chain | 0.103 | 0.125 | 0.411 | 1 |
| Q9H4G4 | GLIPR2 | Golgi-associated plant pathogenesis-related protein 1 | -0.015 | 0.122 | 0.901 | 1 |
| Q9H8L6 | MMRN2 | Multimerin-2 | 0.012 | 0.135 | 0.928 | 1 |
| Q9HDC9 | APMAP | Adipocyte plasma membrane-associated protein | -0.101 | 0.122 | 0.406 | 1 |
| Q9NPH3 | IL1RAP | Interleukin-1 receptor accessory protein | 0.199 | 0.130 | 0.124 | 1 |
| Q9NPR2 | SEMA4B | Semaphorin-4B | -0.153 | 0.126 | 0.223 | 1 |
| Q9NPY3 | CD93 | Complement component C1q receptor | -0.087 | 0.120 | 0.469 | 1 |
| Q9NQ79 | CRTAC1 | Cartilage acidic protein 1 | -0.008 | 0.123 | 0.946 | 1 |
| Q9NTU7 | CBLN4 | Cerebellin-4 | -0.214 | 0.124 | 0.085 | 1 |
| Q9NY15 | STAB1 | Stabilin-1 | -0.060 | 0.134 | 0.656 | 1 |
| Q9NY97 | B3GNT2 | N-acetyllactosaminide beta-1,3-N-acetylglucosaminyltransferase 2 | 0.154 | 0.135 | 0.256 | 1 |
| Q9NZK5 | ADA2 | Adenosine deaminase 2 | -0.190 | 0.130 | 0.145 | 1 |
| Q9NZP8 | C1RL | Complement C1r subcomponent-like protein | 0.170 | 0.132 | 0.199 | 1 |
| Q9UBR2 | CTSZ | Cathepsin Z | 0.080 | 0.127 | 0.529 | 1 |
| Q9UBX1 | CTSF | Cathepsin F | 0.172 | 0.136 | 0.206 | 1 |
| Q9UGM5 | FETUB | Fetuin-B | 0.135 | 0.143 | 0.343 | 1 |
| Q9UHG3 | PCYOX1 | Prenylcysteine oxidase 1 | -0.040 | 0.124 | 0.748 | 1 |
| Q9UJJ9 | GNPTG | N-acetylglucosamine-1-phosphotransferase subunit gamma | 0.107 | 0.125 | 0.394 | 1 |
| Q9UK55 | SERPINA10 | Protein Z-dependent protease inhibitor | 0.262 | 0.147 | 0.074 | 1 |
| Q9ULI3 | HEG1 | Protein HEG homolog 1 | 0.029 | 0.147 | 0.842 | 1 |
| Q9UNN8 | PROCR | Endothelial protein C receptor | -0.178 | 0.133 | 0.182 | 1 |
| Q9UNW1 | MINPP1 | Multiple inositol polyphosphate phosphatase 1 | 0.130 | 0.130 | 0.317 | 1 |
| Q9Y251 | HPSE | Heparanase | 0.015 | 0.128 | 0.907 | 1 |
| Q9Y490 | TLN1 | Talin-1 | -0.160 | 0.128 | 0.214 | 1 |
| Q9Y5C1 | ANGPTL3 | Angiopoietin-related protein 3 | 0.105 | 0.128 | 0.414 | 1 |
| Q9Y5Y7 | LYVE1 | Lymphatic vessel endothelial hyaluronic acid receptor 1 | 0.107 | 0.123 | 0.387 | 1 |
| Q9Y646 | CPQ | Carboxypeptidase Q | 0.098 | 0.131 | 0.452 | 1 |
| Q9Y6R7 | FCGBP | IgGFc-binding protein | 0.041 | 0.138 | 0.767 | 1 |
| Q9Y6Z7 | COLEC10 | Collectin-10 | 0.284 | 0.136 | 0.037 | 1 |
